# Association of CD7⁺CXCR3⁺ CAR T cells with long-term remission in R/R DLBCL

**DOI:** 10.1101/2024.12.06.24318600

**Authors:** Robin Bartolini, Lionel Trueb, Douglas Daoudlarian, Victor Joo, Alessandra Noto, Raphaël Stadelmann, Bernhard Gentner, Craig Fenwick, Matthieu Perreau, Georges Coukos, Giuseppe Pantaleo, Caroline Arber, Michel Obeid

**Affiliations:** Centre Hospitalier Universitaire Vaudois (CHUV), University of Lausanne, Department of Medicine, Immunology and Allergy Service, Rue du Bugnon 46, CH-1011 Lausanne, Switzerland; Centre Hospitalier Universitaire Vaudois (CHUV), University of Lausanne, Department of Oncology, Immuno-Oncology Service, Rue du Bugnon 46, CH-1011 Lausanne, Switzerland; Centre Hospitalier Universitaire Vaudois (CHUV), University of Lausanne, Departments of Oncology and Laboratory Medicine, Hematology Service, Rue du Bugnon 46, CH-1011 Lausanne, Switzerland; Ludwig Institute for Cancer Research, Lausanne Branch, Lausanne, Switzerland; Swiss Cancer Center Léman, Lausanne, Switzerland; AGORA Cancer Research Center, Lausanne, Switzerland

**Keywords:** Anti-CD19 CAR T cell infusion products, immune cell phenotypes, predictive biomarkers, CXCR3, CD7, NKG2D, LAG-3, CD71

## Abstract

**Background.:** CAR T-cell therapy is the standard of care for R/R DLBCL, but more than half of patients fail to achieve long-term remission. Identification of cellular biomarkers in CAR T-cell infusion products (IPs) that predict complete remission beyond six months may guide the development of strategies to improve outcomes.

**Methods.:** IPs from 13 R/R DLBCL patients were analyzed using a 39-marker mass cytometry panel, comparing cell populations between long-term responders (R) and non-responders (NR). Both unsupervised and supervised analyses were performed. Longitudinal blood samples were analyzed for 30 days to track CAR T-cell subpopulation dynamics.

**Results.:** At a median follow-up of 13.5 months, median progression-free survival (PFS) was 13.3 months (95% CI: 9.7-24.3) in R (n=8) versus 3.5 months (95% CI: 0.5-5.4) in NR (n=5).

The HR for PFS was 56.67 (95% CI: 7.3-439.3; P=0.0001). A subset of CD3^+^CXCR3^+^CD7^+^ CAR T-cells found in both CD4^+^ and CD8^+^ populations was significantly enriched in R and expressed higher levels of perforin, granzyme B, and NKG2D (restricted to CD8^+^). NR had more CXCR3^+^CD7^+^LAG3^+^ CAR T-cells. CD3, CD7, CXCR3, and NKG2D cell surface levels were higher in R, whereas LAG3, Ki67, and CD71 were elevated in NR. A predictive cut-off ratio of CD3^+^CXCR3^+^CD7^+^LAG3^+^CAR^+^ T-cells <0.83 and CD3^+^CXCR3^+^CD7^+^NKG2D^+^CAR^+^ T-cells >1.034 yielded a predictive accuracy of 0.92. Serum CXCL9 and CXCL10 levels were not different between groups.

**Conclusions.:** Increased frequency of CAR T-cells expressing CD7, CXCR3 and NKG2D in R versus LAG3 and CD71 in NR emerged as strong correlates of therapeutic outcome.

## Introduction

Despite notable clinical successes, about half of all patients with relapsed and/or refractory diffuse large B-cell lymphoma (R/R DLBCL) do not achieve long-term remissions with currently available CAR T-cell therapy^1^. Predictive factors determining the efficacy remain incompletely understood. The phenotypic composition of infusion products (IPs) is important, as defined T-cell subsets have been associated with enhanced antitumor activity *in vivo*^2^. Patients treated with axicabtagene ciloleucel (axi-cel), achieved higher complete response (CR) rates when their IPs were enriched for CD8 memory T-cell phenotypes^3^. These memory-like T cells are believed to confer sustained proliferative capacity and persistence leading to better clinical outcomes.

Moreover, the expression of inhibitory receptors on CAR-T cells has been linked to therapeutic failure. Human CAR-T cells with intrinsic PD-1 blockade overcome tumor-driven inhibition^4^. Additionally, PD-1 inhibition enhanced the function of CAR-T cells in patient with R/R DLBCL^5^. Notably, exhaustion markers like LAG-3 and TIM-3 on intratumoral CD8⁺ T cells by day 7 post-infusion correlate with poorer outcomes in patients receiving axi-cel^3^. This suggests that regulatory and exhausted T-cell phenotypes may limit the efficacy of CAR T-cell therapy. Similar patterns have been reported in chronic lymphocytic leukemia (CLL), where patients achieving durable remissions following CAR T-cell therapy received infusion products with lower frequencies of CD8⁺ T cells co-expressing inhibitory receptors like PD-1, LAG-3, and TIM-3^6^. Conversely, higher co-expression of these inhibitory receptors correlated with suboptimal responses, highlighting the importance of T-cell fitness and the intricate balance between activation and inhibition in driving successful outcomes^6,7^.

These findings highlight the critical need for a deeper understanding of the phenotypic characteristics that predict CAR T-cell efficacy. By identifying specific CAR T-cell phenotypes associated with successful treatment outcomes, existing IPs and clinical treatment algorithms can potentially be optimized to enhance patient outcomes. Moreover, data from correlative bed-to-bench studies can be exploited for the development of next generation potency-enhanced CAR T-cell products.

To address this knowledge gap, we performed a comprehensive analysis conducted an extensive analysis of the cellular and phenotypic diversity within CAR T-cell infusion products using a 39-marker mass cytometry panel. The objective of this study was to identify predictive markers of durable responses and to improve our comprehension of the factors influencing the long-term efficacy of CAR T-cell therapy in patients with R/R DLBCL.

## METHODS

### Patient Consent, Ethical Approval, and Sample Collection

Participants provided informed consent for the research use of their cells, blood samples, and data through a "consentement général" process, ensuring data confidentiality. Enrollment followed Article 34 of the Swiss Federal Law on Human Research, and the study received approval from the Cantonal Ethics Committee (CER-VD). Biological samples were collected during routine clinical care, with residual CAR T-cells obtained by washing the CAR T-cell infusion bag & line after infusion. Peripheral blood mononuclear cells (PBMCs) were collected from treated patients at multiple time points from week 1 to week 4 post-infusion.

### Mass Cytometry Analysis

A mass cytometry panel comprising 39 markers was developed and validated for this study with the corresponding fluorescently labeled antibody clones from BD Biosciences (**Supplementary Table 1**). The analysis was compared to healthy volunteer data to establish baselines. This panel enabled high-dimensional analysis of cellular phenotypes, offering insights into IP composition and immune cell dynamics in CAR T-cell therapy recipients.

### Study Design and Population

This observational study was conducted at the CHUV’s Immuno-Oncology and Immunology and allergy Service from January 2020 to May 2024. The final analysis included 13 patients with relapsed/refractory (R/R) DLBCL receiving standard of care CAR T-cell therapy and follow up (**Supplementary** Figure 1).

### Oncologic Response Assessment

CAR T-cell therapy efficacy was assessed using PET/CT scans at baseline, and 1-, 3- and 6-months post-infusion.

### Response Definitions

Responders were those with no relapse on PET/CT for at least six months post-infusion. Non-responders either did not respond at the 30-day evaluation or experienced disease progression before the six-month follow-up.

### Statistical Analysis

Statistical analyses were performed using GraphPad Prism 10.1.2 and MATLAB R2023b. ROC curve analysis and clustering were conducted, with optimal cutoffs determined using Youden’s index. Cox proportional hazard model was performed using R and “*survival package*”. Clustering was performed using k-means clustering MATLAB.

## RESULTS

### Patient clinical characteristics

A total of 41 patients treated with CAR T-cell therapy at the University Hospital of Lausanne (CHUV) were identified. CAR-T infusion data were analyzed for 13 patients with R/R DLBCL (study flowchart **Supplementary** Figure 1). Patient characteristics are summarized in **Table** 1. The median follow-up was 13.5 months at the data cut-off date (June 15, 2024). The median patient age was 61 years (range 28-78), with 38% aged >65 years and 77% male (n=10/13). The majority of patients (54%, n=7/13) had stage III or IV disease. Treatments included axicabtagene ciloleucel (axi-cel) (85%, n=11/13) and tisagenlecleucel (tisa-cel) (15%, n=2/13).

Cytokine release syndrome (CRS) was observed in 85% of patients, with 69% exhibiting mild (grades 1-2) and 15% demonstrating severe (grades 3 or higher) CRS, according to the ASTCT Consensus grading system^8^. The most common late (after 30 days) hematologic toxicity (ICAHT) was grade 3 or higher thrombocytopenia (38%) and neutropenia (31%), only grade 3 or higher have been reported in the **Supplementary Table 2**. Regarding CRS treatment, 85% received only tocilizumab (TCZ), 38% received corticosteroids (CS) and 8% received canakinumab (CAN). Four patients were treated in second line, all others in third or higher line, and 31% (n=4/13) had relapsed after autologous stem cell transplantation **(Supplementary Table 2).**

**Table 1.**

|  | N=13 | % within cohort |
| --- | --- | --- |
| <u>Age</u> |  |  |
| Median | 61 |  |
| ≥ 65 years | 6 | 46% |
| <u>Gender</u> |  |  |
| Female | 3 | 23% |
| Male | 10 | 77% |
| <u>Tumor Type</u> |  |  |
| DLBCL | 13 | 100% |
| <u>Initial Stage</u> |  |  |
| I-II | 6 | 46% |
| III-IV | 7 | 54% |
| <u>CAR T-cell type</u> |  |  |
| Axicabtagene ciloleucel | 11 | 85% |
| Tisagenlecleucel | 2 | 15% |
| <u>Cytokines release Syndrome (CRS)</u> |  |  |
| <u>Grading</u> |  |  |
| No CRS | 2 | 15% |
| G1-G2 | 10 | 77% |
| G3-G4 | 1 | 15% |
| G5 | 0 | 0% |
| <u>Immune Effector Cell-Associated Neurotoxicity Syndrome (ICANS)</u> |  |  |
| <u>Grading</u> |  |  |
| No ICANS | 6 | 46% |
| G1-G2 | 3 | 23% |
| G3-G4 | 4 | 31% |
| G5 | 0 | 0% |
| <u>Immune Effector Cell-Associated Hematological Toxicity (ICAHT)</u> |  |  |
| <u>Late (>30 days)</u> |  |  |
| Pancytopenia | 2 | 15% |
| Anemia | 2 | 15% |
| Thrombocytopenia | 5 | 38% |
| Neutropenia | 4 | 31% |
| <u>Immunosuppressive treatment</u> |  |  |
| Corticosteroids (CS) | 5 | 38% |
| Tocilizumab (TCZ) | 11 | 85% |
| Canakinumab (CAN) | 1 | 8% |

### Distinct CAR T-cell populations in infusion products associated with long-term remission

We used a 39-marker panel to identify CAR T-cell populations associated with long-term remission. We first examined the frequencies of total CAR^+^CD4⁺ and CAR^+^CD8⁺ T-cells and the CD4/CD8 ratio but found no significant association with outcomes (**Supplementary** Figure 2). We next extended our analysis to other markers and evaluated their frequencies and expression levels within the total CAR^+^CD3⁺ compartment and the CAR^+^CD3⁺CD4⁺ and CAR^+^CD3⁺CD8⁺ subsets. Among all analyzed markers, we observed significant differential expression of LAG3, Ki67, CD71, CXCR3, CXCR5, CD3, and CD7 between responders (R) and non-responders (NR) (**Figure 1**, **Supplementary** Figure 3). Specifically, IPs of NR contained significantly higher proportions of LAG3^+^, Ki67^+^, and CD71^+^ CAR T-cells in total CD3⁺, CD3⁺CD4⁺, and CD3⁺CD8⁺ compartments, and mean metal intensity (MMI) of these markers was also significantly increased. Conversely, CAR T-cells in IPs of R expressed higher levels of CD3, CD7, CXCR3, and CXCR5 by MMI. Notably, NKG2D expression was higher in R compared to NR but only within the CD8⁺ compartment, consistent with NKG2D’s restriction to CD8⁺ T-cells. Analysis of these markers alone (on the total CAR⁺CD3⁺ compartment) demonstrated significant performance in discriminating between R and NR using receiver operating characteristic (ROC) curve analyses, with area under the curve (AUC) values ranging from 0.85 to 0.975 and p-values from 0.0404 to 0.0054 (**Supplementary** Figure 4)

**Figure 1:**
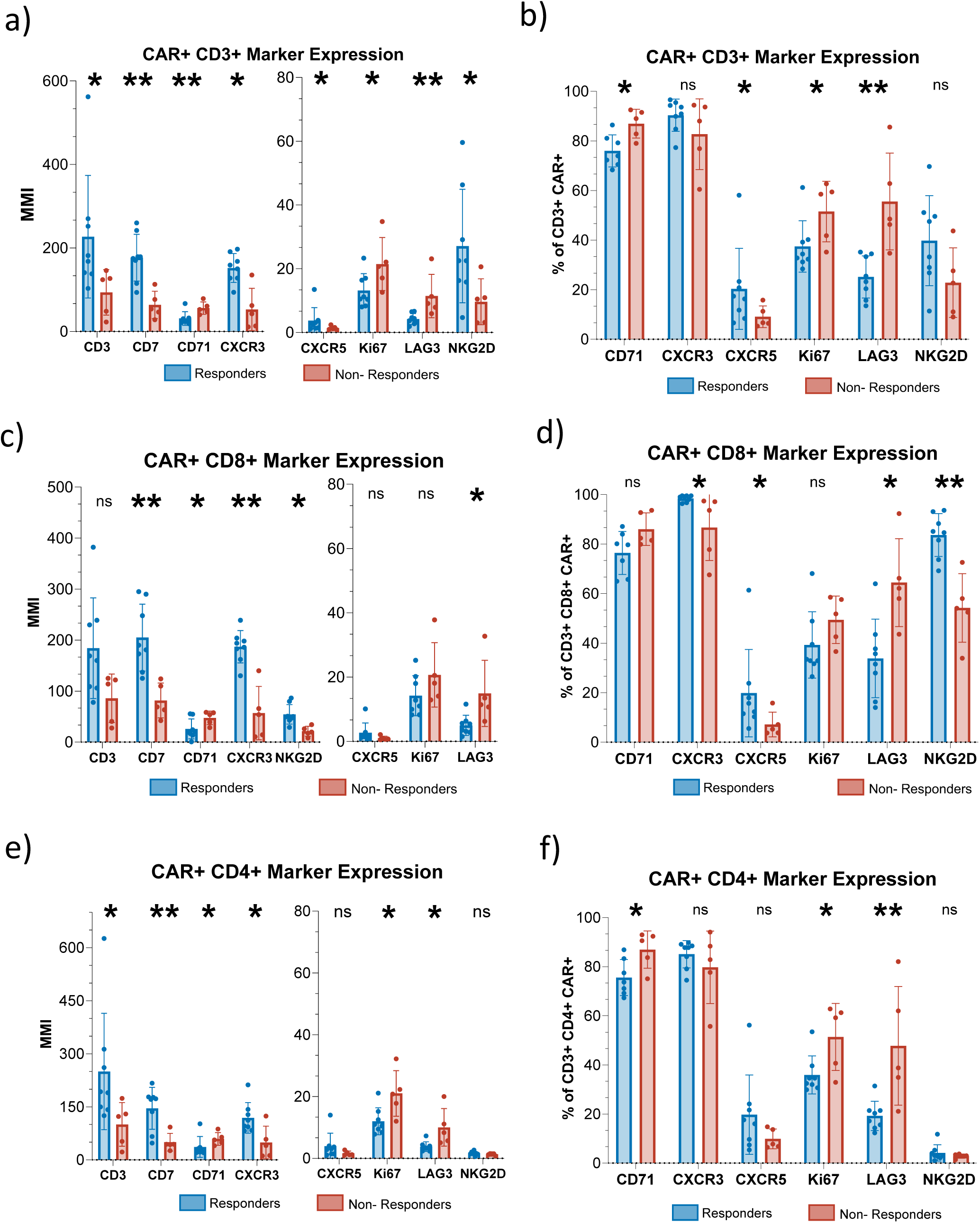
Marker expression in CD3^+^ CAR^+^ T-cells. Expression of CD3, CD7, CD71, CXCR3, CXCR5, Ki67, LAG3 and NKG2D in CD3^+^CAR^+^ T-cells, expressed both as MMI **(a)** and as frequency **(b)** of the parent population. Marker expression was measured in the total CD3^+^ population **(a-b)**, in the cytotoxic **(c-d)** (CD3^+^CD8^+^) and in the helper **(e-f)** (CD3^+^CD4^+^) compartment. Responders=blue, n=8. Non-responders=red, n=5. Statistical significance was determined using the Mann-Whitney U-Test, with significance levels indicated as *p<0.05 and **p<0.01.

### Identification of a responder-enriched CD3⁺CXCR3⁺CD7⁺CAR^+^ T-cell population in infusion products

Next, we investigated whether these identified markers were co-expressed, defining a population that could predict long-term remission. Initially, we used unsupervised UMAP analysis, which divided the CAR^+^CD3⁺ compartment into two clusters: CD4⁺ cells (low NKG2D expression) and CD8⁺ cells. In the CD4⁺ cluster, NR had higher frequencies of populations C4-6 and C4-7, characterized by low CXCR3 and CD7 but high CD71; C4-7 also had elevated Ki67 and LAG3 (**Supplementary** Figure 5). In the CD8⁺ compartment, cluster C8-3 was enriched in R, expressing high CXCR3 and CD7 but low Ki67 and LAG3. Conversely, cluster C8-6, enriched in NR, showed lower CXCR3, CD7, and NKG2D but higher LAG3 compared to C8-3 (**Supplementary** Figure 5). Given the critical importance of CXCR3 and CD7 in defining R and NR, we focused our supervised analysis on CAR^+^CXCR3⁺CD7⁺ double-positive cells. In R, CXCR3⁺CD7⁺ (CD3⁺CAR⁺) cells averaged 85%, compared to 65% in NR. In the CD8⁺ compartment, CXCR3⁺CD7⁺ cells represented 90% of CD8⁺CAR⁺ cells in R versus 75% in NR (**Figure 2A**).

**Figure 2:**
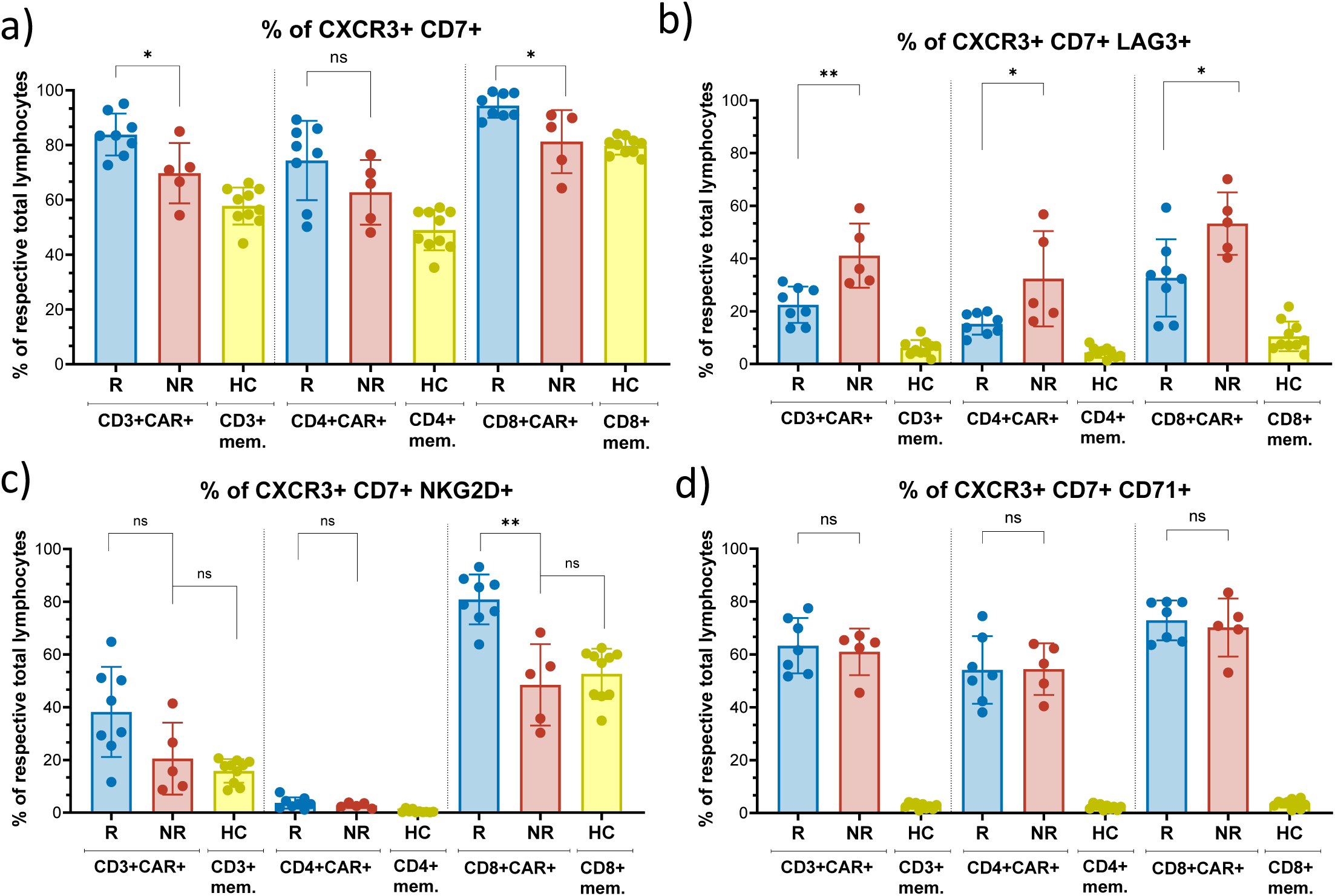
Frequencies of double and triple positive CAR T-cells. **(a)** Frequencies of double- positive CXCR3⁺CD7⁺ cells as a percentage of CAR^+^CD3⁺, CAR⁺CD3⁺CD4⁺, and CAR⁺CD3⁺CD8⁺ cells in responders (blue, n=8) vs. non-responders (red, n=5), **(b-d)** Frequencies of triple-positive CXCR3⁺CD7⁺ (LAG3⁺NKG2D⁺CD71⁺) cells as a percentage of CAR⁺CD3⁺, CAR⁺CD3⁺CD4⁺, and CAR⁺CD3⁺CD8⁺ cells in responders (blue, n=8) vs. non- responders (red, n=5). Memory T-cell frequencies from healthy controls (yellow, n=10) are provided as a baseline reference. Statistical significance determined via Mann-Whitney U-Test, with *p<0.05 and **p<0.01.

### Phenotypic characterization of the CD3⁺CXCR3⁺CD7⁺CAR^+^ T-cell population

We further characterized the CXCR3⁺CD7⁺ double-positive population by comparing co-expression of various other markers to that of CXCR3⁺CD7^-^, CXCR3^-^CD7⁺, and CXCR3⁻CD7⁻ populations. The CXCR3⁻CD7⁻ population expressed the lowest levels of all markers analyzed, except for CD71, which was highly expressed compared to other populations. The CXCR3⁺CD7⁺ cells expressed the highest levels of perforin, granzyme B, and NKG2D, indicating the strongest effector potential (**Supplementary** Figure 6). After identifying this double-positive population in both IPs and longitudinal samples (**Supplementary** Figure 7), we investigated whether other markers on CXCR3⁺CD7⁺ cells were differentially expressed between R and NR. Among all analyzed markers, we again observed significant differences in the expression of CD3, CD7, Ki67, NKG2D, CD71, and LAG3 (**Supplementary** Figure 8). We incorporated these markers one by one into the CXCR3⁺CD7⁺ population. CXCR3⁺CD7⁺LAG3⁺ cells were twice as frequent in the IPs of NR (40% of CD3⁺CAR⁺ cells) compared to R (20%), a difference also observed in the CD4⁺ and CD8⁺ compartments (**Figure 2B**). Conversely, CXCR3⁺CD7⁺NKG2D⁺ cells were significantly more frequent in the CD8⁺ compartment of R, reaching 80% of CD3⁺CD8⁺CAR⁺ T-cells compared to only 45% in NR (**Figure 2C**). No significant differences were observed in the frequency of CXCR3⁺CD7⁺CD71⁺ cells between groups (**Figure 2D**). Interestingly, while the CXCR3⁺CD7⁺ population was also observed in the circulation of healthy individuals, these cells completely lacked CD71 expression, exhibited very low LAG3 levels, and showed NKG2D levels comparable to those observed in NR. NKG2D upregulation in CD3⁺CD8^+^CAR⁺ T-cells seems to be exclusive to R. This highlights profound phenotypic differences between normal memory T-cells and CAR T-cells, suggesting that the CXCR3⁺CD7⁺CAR^+^ population in responders is uniquely endowed with cytotoxic capabilities essential for effective antitumor responses.

### The predictive value of marker ratios for six-month progression-free survival

Given the established correlation between specific markers and patient survival, we sought to determine whether the balance between positive and negative subpopulations for CD71, LAG3, and NKG2D within the CXCR3⁺CD7⁺ subset influences the prediction of treatment response. We evaluated the ratios of CD71, LAG3, and NKG2D MMI within the CXCR3⁺CD7⁺ population across all CAR^+^ T-cells as well as within CAR^+^CD4⁺ and CAR^+^CD8⁺ subsets, to assess their predictive performance. The analysis demonstrated that all ratios—except for NKG2D in CD3⁺ cells and CD71⁺ in CAR^+^CD8⁺ cells—showed statistically significant differences between R and NR. Optimal cutoff values were identified using the Youden index and utilized to reclassify patient responses using forementioned ratios, all markers except NKG2D in CD3 and CD71 in CAR^+^CD8^+^ had ROC that were predictive, with accuracies ranging from 0.77 to 0.92. Notably, LAG3 positivity and NKG2D negativity within the CXCR3⁺CD7⁺ population were linked to poorer PFS outcomes (**Figure 3**). We then examined the expression levels of previously identified significant markers—CXCR3, CD7, NKG2D, LAG3, and Ki67—on various populations based on CD3⁺, CD4⁺, CD8⁺, and CXCR3⁺CD7⁺ subsets to determine whether these levels could predict long-term remission. Our findings revealed that all marker MMIs, except for Ki67 in the CD4⁺CXCR3⁺CD7⁺ and CD8⁺CXCR3⁺CD7⁺ subsets, demonstrated substantial capacity for differentiating between R and NR through ROC curve analyses. The AUC values ranged from 0.85 to 1, with p-values from 0.0034 to 0.0415 (**Supplementary** Figure 9). Notably, CD7 MMI exhibited optimal performance, reaching an AUC of 1 in CD8⁺ cells (p = 0.0034). A comparable outcome was obtained using frequencies of marker expression (**Supplementary** Figure 10). Using the ROC based approach on both MMI (**Supplementary** Figure 11) and frequencies (**Supplementary** Figure 12), KM analyses revealed that most markers significantly predicted six-month PFS. We calculated accuracy, negative predictive value (NPV), positive predictive value (PPV), sensitivity, and specificity for these predicted PFS values. The most effective predictors were CD7 MMI in both CD8⁺ and CD8⁺CXCR3⁺CD7⁺ CAR-T cells. Other markers demonstrated satisfactory accuracy, ranging from 0.77 to 0.92. Notably, several markers exhibited PPV or NPV values of 1, indicating a robust correlation between their expression and six-month PFS.

**Figure 3:**
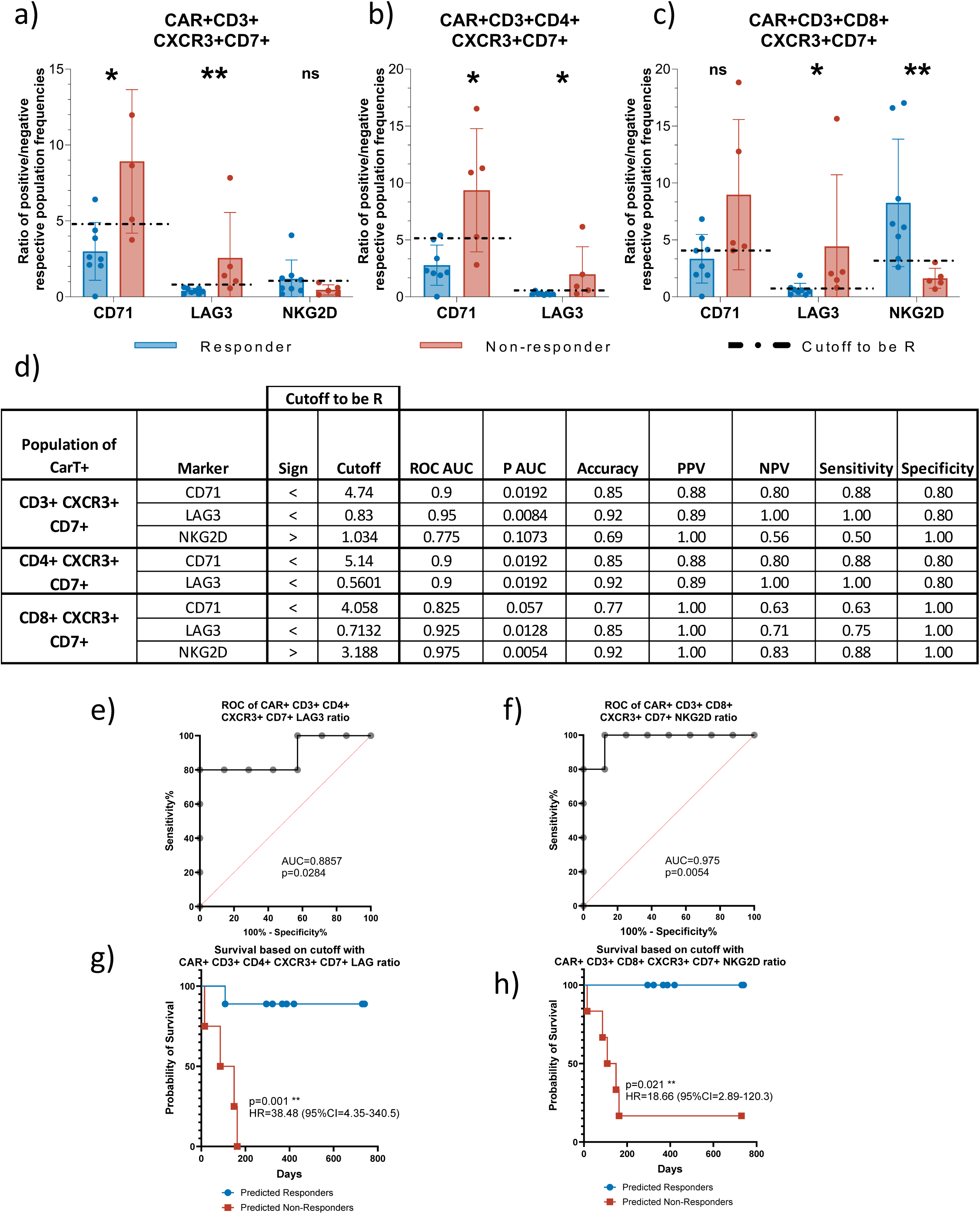
Marker ratios as predictive tools for six-month PFS. **(a)** Ratios of positive/negative populations for CD71, LAG3, and NKG2D in CAR⁺CXCR3⁺CD7⁺CD3⁺ (**a**), CD3⁺CD4⁺ (**b**), and CD3⁺CD8⁺ (**c**) cells in responders (blue, n=8) vs. non-responders (red, n=5). NKG2D in CD4⁺ is excluded due to lack of expression. Statistical significance determined by Mann-Whitney U-Test (*p<0.05, **p<0.01), **(d)** Summary table of predictive power using marker ratios calculated with optimal cutoff values determined via the Youden index. New predicted PFS accuracies, positive predictive value (PPV), negative predictive value (NPV), sensitivities, and specificities are reported. **(e-f)** ROC and AUC for the ratio of LAG3 in CD4+ and NKG2D in CD8+ and **(g-h)** association of the respective ratio cutoff to progression free survival.

### Clustering analysis for predicting PFS

To further evaluate the predictive potential of specific CAR-T subpopulation frequencies and marker expression levels independently of the known PFS, we assessed whether CXCR3 and CD7 MMI in total CD3⁺CAR^+^, CD3^+^CD4⁺CAR^+^, or CD3^+^CD8⁺CAR^+^ T cells could discriminate between R and NR. One-dimensional k-means clustering was used to categorize MMI expression into two clusters. The predictive accuracy of these clusters for six-month PFS was then evaluated using KM curves. This approach yielded statistically significant results, with p-values ranging from 0.001 to 0.016 for all clusters except CD8⁺CD7⁺ MMI (p = 0.1646) (**Supplementary** Figure 13a–f). These findings indicate that elevated expression levels of CXCR3 and CD7 are associated with enhanced PFS in most T-cell subsets examined. We also applied the same clustering approach to differentiate between R and NR based on the simultaneous expression of multiple markers—CXCR3, CD7, CD71, NKG2D, LAG3, and Ki67 MMI—on the CD3⁺CXCR3⁺CD7⁺ population (**Supplementary** Figure 14a). The silhouette value approach determined an optimal two-cluster solution (**Supplementary** Figure 14b), identifying two distinct clusters: Cluster 1 consisted of four NR, while Cluster 2 included all eight R and one NR. t-SNE visualization showed that the four NR in Cluster 1 clustered closely, whereas the one NR in Cluster 2 was closer to the R (**Supplementary** Figure 14c). This underscores the notable phenotypic distinction of infused CAR-T cells between most NR and R.

### Correlation analysis of marker expression

The clustering analysis demonstrated clear distinctions of CAR T-cell phenotypes independent of infusion product type; NR clustered together regardless of the presence of a CD28 or 4-1BB co-stimulatory domain. This pattern suggests that certain markers are correlated, forming two distinct phenotypes associated with response status. The NR phenotype is characterized by high expression of LAG3 and CD71 with low levels of CXCR3 and CD7, while the R phenotype exhibits high CXCR3 and CD7 with low LAG3 and CD71.

A correlation matrix of all 39 markers expression across CAR^+^CD3⁺, CD4⁺, and CD8⁺ compartments revealed intriguing population dynamics. Specifically, positive correlations were observed between CD7 and CXCR3, and between CD71 and LAG3 expressing CAR T-cells. Negative correlations were noted between CXCR3 and LAG3, CD71 and NKG2D, and NKG2D and LAG3 expression (**Supplementary** Figures 15–17). These negative correlations imply mutual exclusivity among certain markers—for example, high CXCR3 expression coincided with low LAG3 expression. A positive correlation was observed between the frequencies of LAG3 and Ki67-expressing CAR T-cells across all T cell subsets, including total CD3^+^, CD4^+^, and CD8^+^ populations. Furthermore, within the CD8^+^ CAR T-cell subset, a significant positive correlation was identified between the frequencies of CXCR3 and NKG2D- expressing CAR T-cells **(Supplementary** Figures 18-20).

### Longitudinal analysis of the CXCR3⁺CD7⁺ population post-infusion

After characterizing the CXCR3⁺CD7⁺ population in IPs of both R and NR, and identifying markers linked to long-term remission, we evaluated the population dynamics in circulation up to 30 days post-infusion. Our focus was on the frequency of these cells and alterations in marker expression. At baseline, all patients were lymphopenic. Between Day 3 and 7 post-infusion, CD3^+^ cell frequencies increased in both R and NR. However, while there was no difference in the expansion of CAR⁻ T-cells, we observed a substantial increase in CAR⁺ T-cells only in R (**Supplementary** Figure 21). Specifically, this expansion was driven by CXCR3⁺CD7⁺ CAR⁺ T-cells, which reached 12.0% of total CD45⁺ cells at Days 7, compared to 2.7% in NR (**Figure 4a**). In R, this represented a tenfold increase compared to Day 3, whereas expansion was minimal in NR. In contrast, the frequencies of CXCR3⁻CD7⁺, CXCR3⁺CD7⁻, and double-negative (CXCR3⁻CD7⁻) CAR⁺ T-cells remained low and were similar between R and NR (**Supplementary** Figure 22). Between Days 3 and 10, CXCR3⁺CD7⁺ CAR⁺ T-cells in R showed significant increases in CXCR3 and Ki67 expression levels, suggesting enhanced proliferation and activation. These increases were not observed in NR (**Figure 4b**). On Day 7, CXCR3⁺CD7⁺ CAR⁺ T-cells in R exhibited increased expression of CXCR3 and Ki67 compared to NR, while LAG3 levels in NR were non significantly elevated relative to R (**Figure 4 c-e**). Circulating levels of CXCR3-binding chemokines, CXCL9 and CXCL10, were similarly elevated in both R and NR and did not increase during the expansion phase (**Figures 4f–g**). These findings indicate that enhanced activation and proliferation of CXCR3⁺CD7⁺ CAR⁺ T-cells are essential for a favorable therapeutic outcome and occur independently of circulating CXCR3-binding chemokine levels.

**Figure 4:**
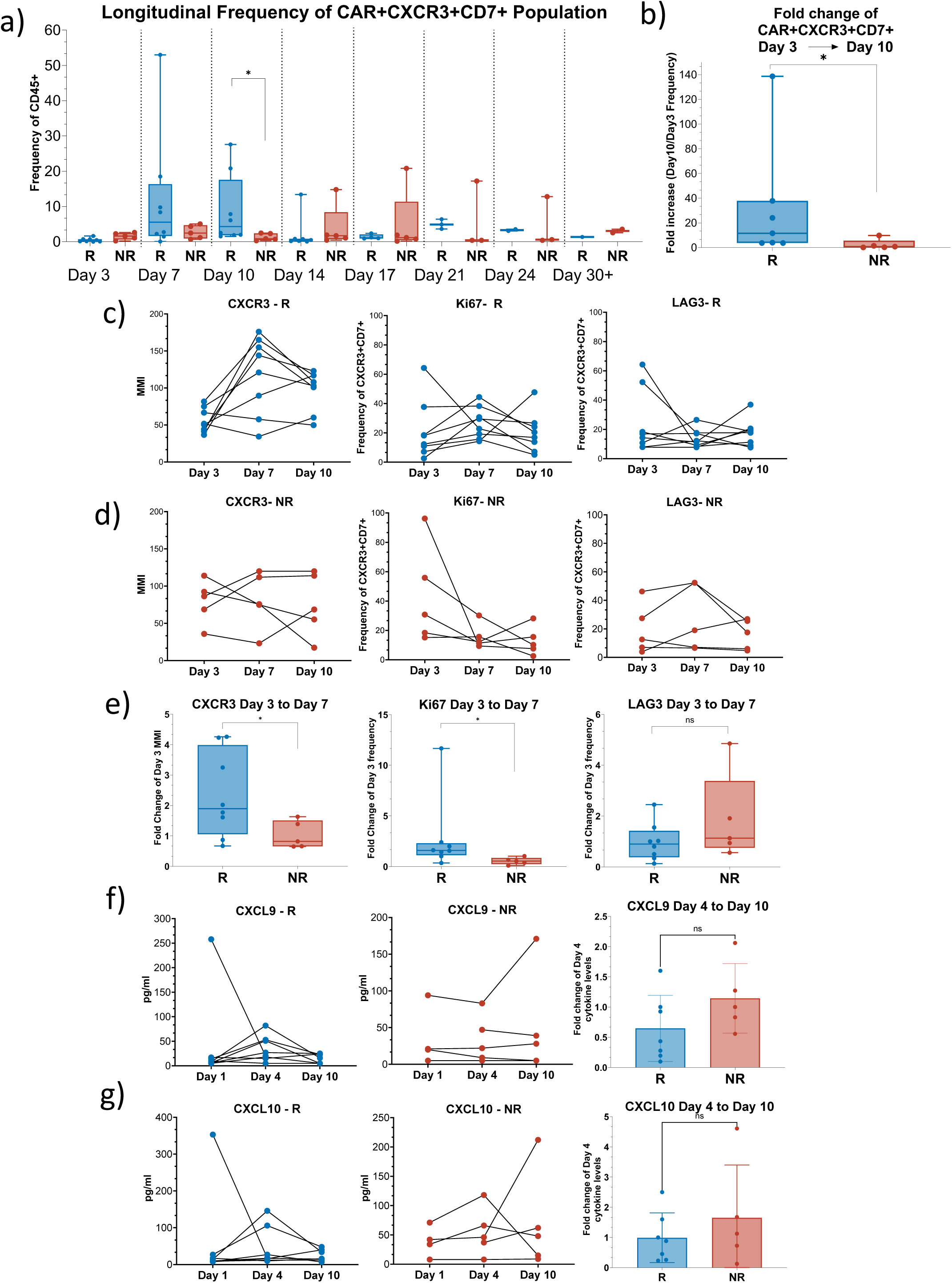
Longitudinal analysis of CXCR3⁺CD7⁺ CD3⁺CAR⁺ T-cell frequency and marker expression. **(a)** Frequency of CAR^+^CD3⁺CXCR3⁺CD7⁺ cells in patient circulation (% of total CD45⁺ events) from Day 3 to Day 30 post-infusion (responders in blue, non-responders in red), **(b)** Fold change in frequency of CXCR3⁺CD7⁺ (CAR⁺CD3⁺, % of total CD45⁺ events) from Day 3 to Day 10, comparing responders (n=8, blue) to non-responders (n=5, red) **(c-d)** Paired analysis of CXCR3 (as MMI), Ki67 and LAG3 expression fluctuations (as frequency) in the CXCR3⁺CD7⁺ population in responders (blue) and non-responders (red) between Days 3 and 10. **(e)** Associated graphs showing fold change in CXCR3, Ki67 and LAG3 MMI between Days 3 and 7. **(f-g)** Levels of CXCR3-binding chemokines CXCL9 and CXCL10 in circulation of responders (blue) and non-responders (red) at Day 0 (infusion), Day 4, and Day 10 post- infusion, with associated graphs showing fold changes in cytokine levels from Day 4 to Day 10. Statistical significance was assessed using the Mann-Whitney U-Test, with significance levels indicated as *p<0.05 and **p<0.01. The line shown within each box plot represents the median.

From Day 10 onwards, we observed a significant elevation of perforin levels in CXCR3⁺CD7⁺ CD3⁺ CAR⁺ T-cells in R. This increase was not due to changes in CD4⁺ or CD8⁺ frequencies, as proportions of CD4⁺CXCR3⁺CD7⁺ and CD8⁺CXCR3⁺CD7⁺ cells remained similar between R and NR (**Supplementary** Figure 23). Apart from perforin, CXCR3, LAG3, and Ki67, no other markers showed a comparable expression pattern over time. These findings suggest that in R, the CXCR3⁺CD7⁺ CAR⁺ T-cell population not only expands significantly post-infusion, but also shows enhanced proliferative and cytotoxic capabilities. The absence of similar changes in NR underscores the potential role of dynamic shifts in the CXCR3⁺CD7⁺ population in mediating effective antitumor responses and achieving long-term remission.

## Discussion

Our study provides novel insights into the phenotypic characteristics of CAR T-cells in standard of care IPs that are associated with long-term remission in patients with R/R DLBCL by identifying specific markers and cell populations that distinguish R from NR. These findings could have substantial implications for predicting treatment outcomes, optimizing CAR-T cell manufacturing processes, and ultimately improving patient outcomes. We observed that the overall CD4/CD8 ratio did not correlate with outcomes, aligning with previous reports suggesting that quantitative global measures of T cell lineage subset content alone may not sufficiently predict clinical responses to CAR T-cell therapy^9^. This prompted a deeper exploration into phenotypic and functional characteristics of CAR T-cell populations. Here, we report differential expression of activation and exhaustion markers between R and NR. NR had higher levels of LAG3, Ki67, and CD71, indicating a state of enhanced activation and potential early exhaustion of CAR-T cells prior to infusion. LAG3 is an inhibitory receptor associated with T cell exhaustion and impaired effector function^10^. It was reported that the disruption of LAG3 in CAR T cells enhanced their anti-tumor activity by improving their proliferation and persistence, leading to improved tumor eradication in preclinical models^11,12^, highlighting the potential of targeting LAG3 as a tool to improve CAR T-cell efficacy, either by direct modification of CAR-T cells or by combining CAR T-cell therapy with LAG3 inhibitors. Elevated levels of Ki67 and CD71 indicate active proliferation and increased metabolic demand, leading to replicative senescence and reduced persistence after infusion^13^, which may contribute to suboptimal clinical outcomes.

In contrast, R expressed higher levels of CD3, CD7, CXCR3 and NKG2D. CXCR3 is involved in T-cell trafficking and homing to sites of inflammation and malignancy^14,15^, suggesting enhanced migratory capabilities of CAR T cells in R. NKG2D is an activating receptor recognizing stress-induced ligands on tumor cells, contributing to increased cytotoxicity^16,17^. It was reported that NKG2D-expressing CAR T-cell therapies, either alone or in combination with other treatments like IL-15 or radiotherapy, may offer an effective approach to enhancing CAR T-cell therapy’s efficiency against various cancers^18^. The elevated expression of these receptors on CAR T cells suggests that CAR T cells from R are likely endowed with enhanced migratory and effector functions, which facilitates effective tumor localization and eradication. Notably, the CXCR3⁺CD7⁺ double-positive population, enriched in R, was characterized by the strongest effector potential with detection of high perforin, granzyme B, and NKG2D levels. In contrast, IPs of NRs contained a higher frequency of CXCR3⁺CD7⁺LAG3⁺CAR^+^ T cells, suggesting CAR-T cell susceptibility to inhibitory signals that might limit efficacy even within potentially beneficial subsets.

Our results demonstrate that certain markers—such as CD7 MMI in CD8⁺ cells—were strongly predictive for six-month progression-free survival. ROC analyses and clustering approaches reinforced the utility of these markers for distinguishing R from NR. R experienced a significant expansion of CXCR3⁺CD7⁺CAR⁺ T cells post-infusion, accompanied by increased perforin, Ki67 and CXCR3 expression, indicating enhanced proliferation and activation, necessary prerequisites for achieving sustained remission^19^. The absence of these features in NR, along with upregulated LAG3, may contribute to poorer outcomes.

Implementing biomarker assessments, such as incorporating key markers in flow cytometry panels, could improve CAR T-cell product evaluation, inform real-time clinical decisions, and aid in patient stratification. The identified markers could also guide modifications to the CAR T-cell manufacturing process—such as enriching for favorable markers (e.g., CXCR3⁺CD7⁺NKG2D⁺) and depleting those associated with exhaustion (e.g., LAG3⁺). Additional strategies may include engineering CAR T cells to downregulate inhibitory receptors such as LAG3 or overexpress activating receptors like NKG2D, thereby enhancing antitumor activity. The administration of checkpoint inhibitors targeting LAG3 at an early stage may prove beneficial for patients with suboptimal expansion or an elevated level of inhibitory receptors following infusion. These questions require assessment in carefully designed clinical trials.

However, our study’s relatively small sample size may limit the generalizability of the findings. Future studies involving larger, multicenter cohorts are needed to validate these biomarkers. Incorporating functional assays alongside phenotypic analysis would also provide a more comprehensive understanding of their roles in mediating therapeutic effects. The immune microenvironment and patient-specific factors may also influence CAR T-cell efficacy, suggesting that a multifaceted approach is needed to fully optimize therapy.

This study underscores the value of phenotypic characterization for predicting clinical outcomes in CAR T-cell therapy. By integrating these biomarkers into clinical protocols, we can enhance patient selection, tailor manufacturing processes, and implement targeted interventions to improve the efficacy of CAR T-cell treatments. Future research should focus on validating and operationalizing these markers to maximize their potential in personalized medicine.

## Supporting information

Supplementary figure merged

## Data Availability

The datasets supporting the results of this study are not publicly available. Requests for access to the dataset will be granted upon reasonable request to the principal investigator. Study data will be managed, stored, shared, and archived according to CHUV standard operating procedures to ensure the continued quality, integrity, and utility of the data.

## Acknowledgements

This work was supported by the strategic plan of the CHUV. We would like to express our gratitude to all the patients who generously contributed their time and samples for this project.

## Author contributions

MO designed the study and supervised the analysis. MO, RB and DD drafted the manuscript. MO, RB and DD had full access to all data in the study and take responsibility for its integrity and accuracy. MO, DD, RB, LT and CA collected the data. AN, CF, MP, GP and MO developed the CAR T-cell panel. RB, DD, VJ, CA, BG, LT and MO analyzed, interpreted and discussed the data. LT, RS and CA recruited patients and supervised the clinical treatments. MO participated in the clinical treatments. RB, DD, VJ and MO prepared the figures. GC and GP participated in the scientific discussion. The manuscript was reviewed and approved by all authors before submission.

## Graphical Abstract

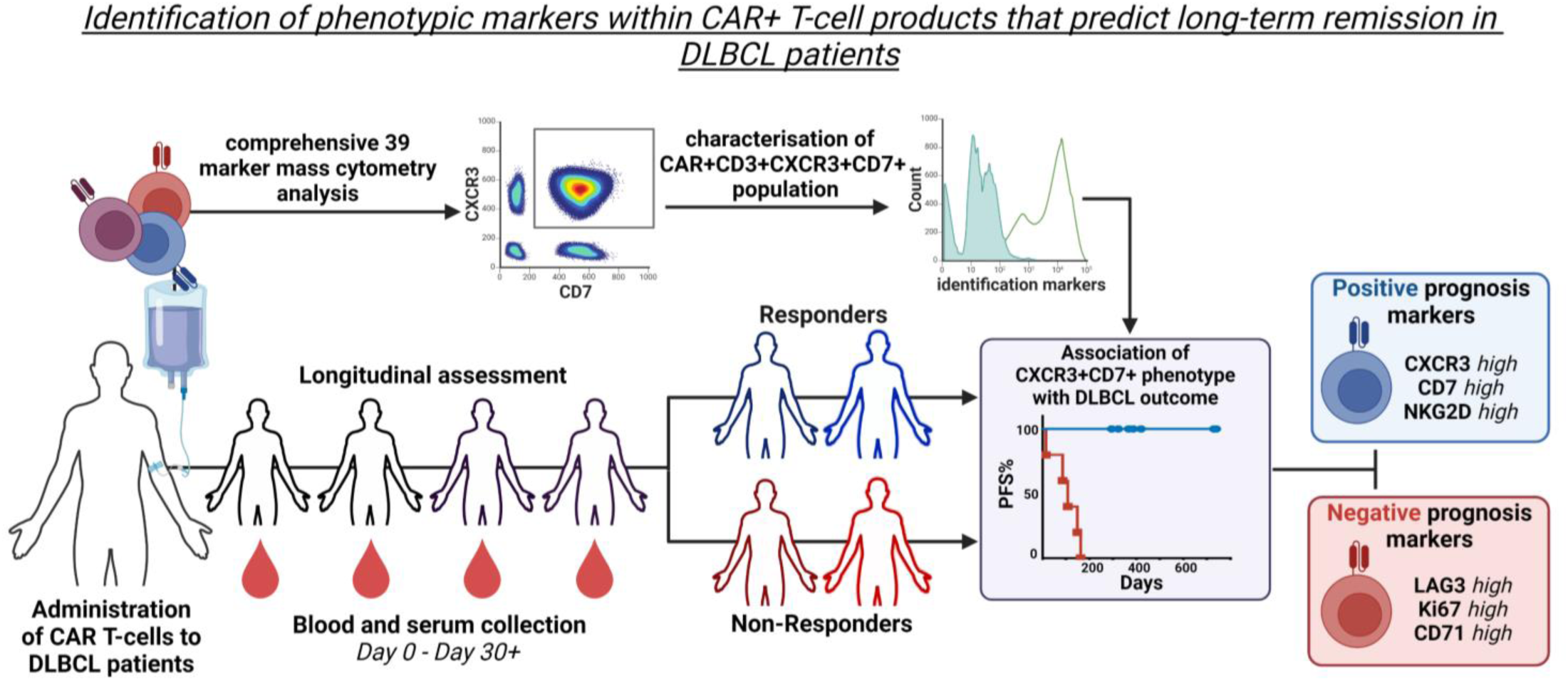

