## Supplementary figure merged for "Association of CD7⁺CXCR3⁺ CAR T cells with long-term remission in R/R DLBCL"

This appendix has been provided by the authors to give readers additional information about the work

Lausanne Center for Immuno-Oncology Toxicities LCIT

Centre Hospitalier Universitaire Vaudois (CHUV)

### Supplementary Methods

#### Immune Profiling of CAR T-Cell Phenotype

Patient blood cells were processed using a standardized whole blood staining protocol[1], with the same procedure applied to cells recovered from the infusion bag (except for the 15-minute RBC lysis step, which was replaced by a simple PBS wash). Briefly, 200 µL of blood containing immune cells were incubated for 30 minutes at 4°C with a 50 µL antibody cocktail of metal-conjugated antibodies targeting CD8, CD4, CCR4, CD127, CCR6, CXCR3, CCR7, CXCR5, and CD45 (Standard BioTools). The cells were then washed with PBS (Laboratorium Dr. G. Bichsel AG) and fixed with 2.4% PFA (Thermo Fisher Scientific) for 10 minutes at room temperature. Cells were lysed for 15 minutes at room temperature using 4 mL of Bulklysis solution (Cytognos), followed by a wash to remove the Bulklysis solution. After another PBS wash, the cells were incubated for 30 minutes at room temperature with a 50 µL antibody cocktail of metal-conjugated monoclonal antibodies targeting CD3, CD19, ICOS, TIGIT, OX40, PD1, CD95, CD62L, CD27, CD25, CD45RO, NKG2D, CD38, CD66b, TIM-3, CD45RA, LAG3, HLA-DR, CD56, CD57, CD71, and CD16. Cells were then permeabilized for 30 minutes at 4°C using 1 mL of Fix/Permeabilization buffer from the Foxp3 Fixation/Permeabilization Kit (eBioscience). Following a wash with Permeabilization Wash buffer, the cells were stained for 30 minutes at 4°C with a 50 µL antibody cocktail of metal-conjugated monoclonal antibodies against Ki67, BCL2, Perforin, and Granzyme B. The cells were washed, and total cells were identified by DNA intercalation using 1 µM Cell-ID Intercalator (Standard BioTools) in 2% PFA at 4°C overnight. Labeled samples were acquired using the HELIOS CyTOF system (Standard BioTools), and FCS files were normalized to EQ Four Element Calibration Beads using CyTOF software. Gates were determines using Fluorescent Minus One (FMO) controls. A complete list of antibodies, including clones, epitopes, and dilutions, can be found below.

| Antibody List | | | | | |
|---|---|---|---|---|---|
| **Metal isotope** | **Target** | **Function** | **Clone** | **Distributor** | **Dilution** |
| **89 Y** | CD45 | Pan-luekocyte marker | HI30 | Biolegend | 1:100 |
| **110 Cd** | CCR4 | Chemokine homing to skin | L291H4 | Biolegend | 1:200 |
| **113 In** | CD8 | Cytotoxic T lymphocyte marker | RPA-T8 | Biolegend | 1:200 |
| **115 In** | CD4 | Helper T lymphocyte marker | RPA-T4 | Biolegend | 1:400 |
| **138 Ba** | Beads | Calibration and normalisation | 291978 | Fluidigm | 0.1x |
| **141 Pr** | CCR6 | Chemokine homing to mucosa | 11A9 | BD Biosciences | 1:200 |
| **142 Nd** | CD19 | B-cell marker | HIB19 | Fluidigm/DVS | 1:142.86 |
| **143 Nd** | ICOS (CD278) | Co-stimulatory molecule | C398.4A | Biolegend | 1:111.11 |
| **144 Nd** | TIGIT | Immune checkpoint receptor | MBSA43 | Fluidigm/DVS | 1:142.86 |
| **145 Nd** | CART | Chimeric Antigen Receptor | REA1298/REA746 | Miltenyi | 1:100 |
| **147 Sm** | CD7 | T-cell marker | CD7-6B7 | Biolegend | 1:100 |
| **149 Sm** | CD127 | IL-7 receptor alpha chain | A019D5 | Fluidigm/DVS | 1:142.86 |
| **150 Nd** | OX40 (CD134) | Co-stimulatory molecule | ACT35 | Fluidigm/DVS | 1:100 |
| **151 Eu** | PD-1 | Immune checkpoint receptor | EH12.2H7 | Biolegend | 1:400 |
| **152 Sm** | CD95 | Fas, memory T marker | DX2 | ThermoFisher | 1:142.86 |
| **153 Eu** | CD62L | L-selectin, adhesion molecule | DREG-56 | Fluidigm/DVS | 1:333.33 |
| **154 Sm** | CD3 | T-cell marker | UCHT1 | BD Biosciences | 1:100 |
| **155 Gd** | CD27 | Co-stimulatory receptor | L128 | Fluidigm/DVS | 1:200 |
| **158 Gd** | CD25 | IL-2 receptor alpha chain | M-A251 | Biolegend | 1:333.33 |
| **159 Tb** | CCR7 | Chemokine homing to lymphoid tissues | G043H7 | Biolegend | 1:100 |
| **160 Gd** | CD14 | Monocyte marker | M5E2 | Fluidigm/DVS | 1:105.26 |
| **161 Dy** | Ki67 | Nuclear proliferation marker | Ki67 | Biolegend | 1:142.86 |
| **162 Dy** | CD69 | Early activation marker | FN50 | Biolegend | 1:200 |
| **163 Dy** | CXCR3 | chemokine homing to inflamed tissues | G025H7 | Biolegend | 1:100 |
| **164 Dy** | CXCR5 | chemokine homing to follicles | J252D4 | Biolegend | 1:100 |
| **165 Ho** | CD45RO | memory isoform of CD45 | UCHL1 | Fluidigm/DVS | 1:200 |
| **166 Er** | NKG2D | activating receptor | ON72 | Fluidigm/DVS | 1:166.67 |
| **167 Er** | CD38 | activation and proliferation marker | HIT2 | Biolegend | 1:250 |
| **168 Er** | CD66b | Neutrophil marker | G10F5 | BD Biosciences | 1:100 |
| **169 Tm** | TIM-3 | Immune checkpoint receptor | PCH101 | Fluidigm/DVS | 1:100 |
| **170 Er** | CD45RA | naive isoform of CD45 | HI100 | Fluidigm/DVS | 1:166.67 |
| **171 Yb** | Granzyme B | secreted cytotoxic effector molecule | GB11 | Fluidigm/DVS | 1:333.33 |
| **172 Yb** | LAG3 | Immune checkpoint receptor | 874501 | R&D Systems | 1:100 |
| **173 Yb** | BCL2 | anti-apoptotic protein | 100 | Biolegend | 1:200 |
| **174 Yb** | HLADR | antigen presentation and T-cell activation | L243 | Fluidigm/DVS | 1:333.33 |
| **175 Lu** | Perforin | secreted cytotoxic effector molecule | B-D48 | Fluidigm/DVS | 1:166.667 |
| **176 Yb** | CD56 | NK/NKT cell marker | HCD56 | Fluidigm/DVS | 1:142.86 |
| **Ir 191/193** | DNA | DNA intercalator | Cell-ID | Standard BioTools | 1µM |
| **194 Pt** | CD57 | marker of terminal differentation | NK-1 | Biolegend | 1:142.86 |
| **196 Pt** | CD71 | transferrin receptor, proliferation marker | AC102 | Fluidigm/DVS | 1:166.67 |
| **209 Bi** | CD16 | Fc receptor, monoyte marker | 3G8 | Fluidigm/DVS | 1:333.33 |

**Supplementary Table 1- Detailed antibody list for mass Cytometry**

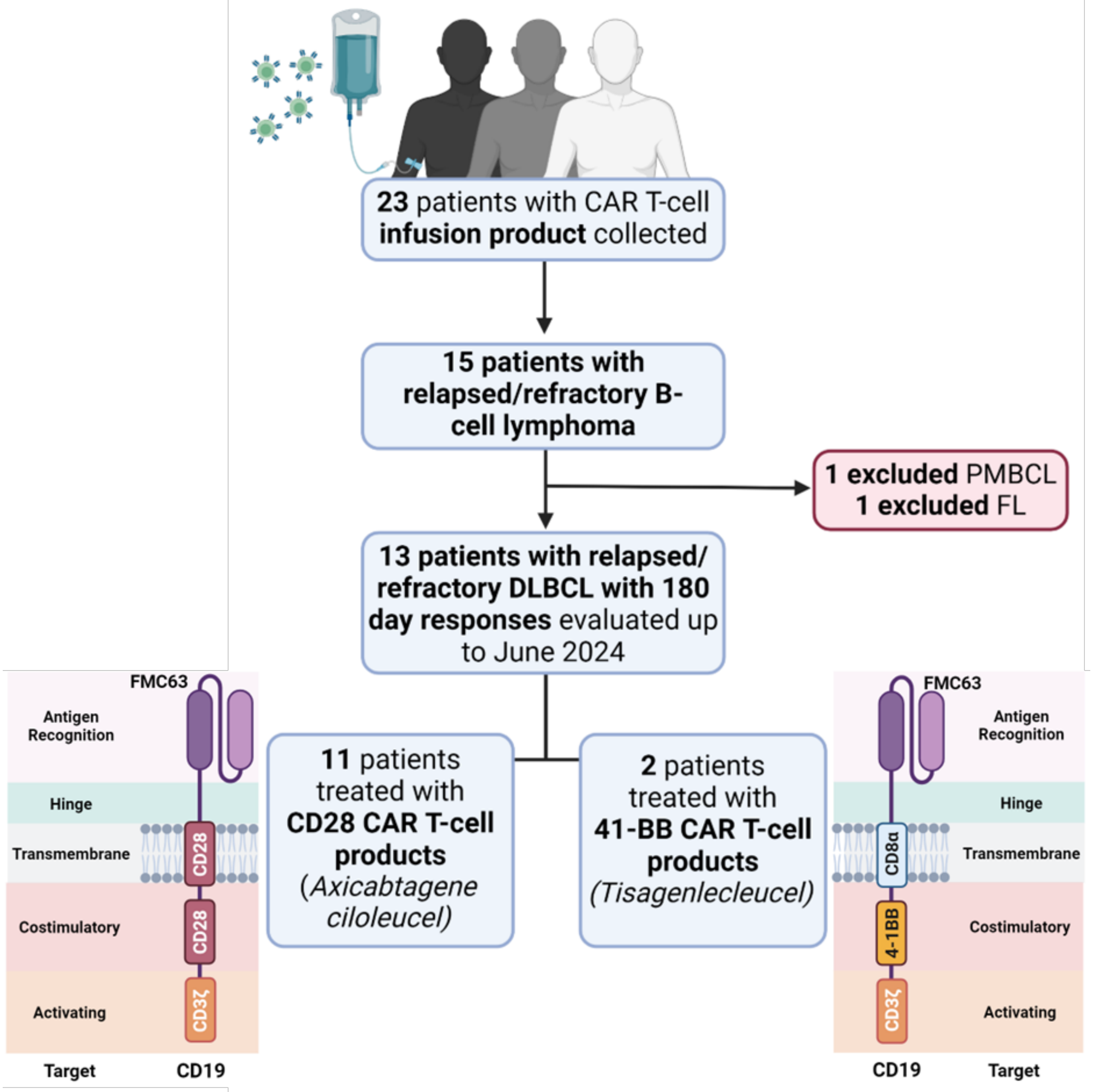

**Figure S1- The flowchart of the study.**

| ID | 180 Days Response | Genetic and molecular features or characteristics | CAR-T Treatment line | Previous Treatment lines | Bridging | Lymphodepleting Chemotherapy |
|---|---|---|---|---|---|---|
| 1 | Non-responder | CD5+, EBV- BCL2+ MYC+, IGH/MYC rearrangement | 3L | 1- MISTRAL-1 and BEAM + ASCT<br>2- R-CHOP | Radiotherapy | Flubarabine + Cyclophosphamide |
| 2 | Responder | CD30-, EBV-, No BCL2/BCl6/MYC Gene rearrangement | 2L | 1- R-CHOP | Radiotherapy | Flubarabine + Cyclophosphamide |
| 3 | Responder | BCL2 with gene rearrangement and hypermutation, MYC+. No gene rearrangement for BCL6 and MYC | 3L | 1- R-CHOP (2 cycles), then R-MTX-ARAC (2 cycles), followed by BCNU-TTP + ASCT<br>2- R-MTX | R-MTX | Flubarabine + Cyclophosphamide |
| 4 | Responder | BCL+, MYC+/-, P53+/-, Mib-1 70%. No gene rearrangement for MYC/BCL2/BCL6 | 2L | 1- R-CHOP | R-ICE | Flubarabine + Cyclophosphamide |
| 5 | Responder | BCL6+ with gene rearrangement. No gene rearrangement for BCL2/MYC | 2L | 1- R-CHOP | R-ICE | Flubarabine + Cyclophosphamide |
| 6 | Non-responder | BCL2+, MYC-, p53+ without mutation | 3L | 1- R-CHOP (2 cycles), intensification with R-ICE (3 cycles) + Ibrutinib, followed by BEAM+ASCT<br>2- Ibrutinib+Venetoclax | Ibrutinib-Venetoclax | Flubarabine + Cyclophosphamide |
| 7 | Non-responder | BCL6 + MYC gene rearrangement | 3L | 1- R-CHOP<br>2- R-ICE | R-DHAOx | Flubarabine + Cyclophosphamide |
| 8 | Responder | BCL2+, MYC+, with BCL2 gene rearrangement | 3L | 1- R-CHOP<br>2- R-ICE, followed by BEAM-ASCT | Radiotherapy | Flubarabine + Cyclophosphamide |
| 9 | Responder | CD19+, BCL2+ MYC+. No gene rearrangement for BCL2/BCL6/MYC. LOH for TP53, and SF3B1 mutation | 8L | *1- Bendamustine+Rituximab*<br>*2- FCR*<br>*3- Venetoclax+Ibruitinib*<br>*4- Venetoclax+Rituximab*<br>*5- Venetoclax+Ibrutinib*<br>*6- RIC allogeneic MMUD-HSCT*<br>7 - ibrutinib R-CHOP | Ibrutinib - R-CHOP | Flubarabine + Cyclophosphamide |
| 10 | Responder | BCL2+, CD30,MYC+. No gene rearrangement for BCL2/BCL6/MYC | 3rd | 1- R-CHOP<br>2- R-ICE | Ibrutinib | Flubarabine + Cyclophosphamide |
| 11 | Non-responder | CD20+/-, Kappa+, MYC+, BCL2+, P53+/-, CD30- | 5L | 1- R-CHOP (1 cycle), then R-DA-EPOCH (6 cycles) and HD-MTX (2 cycles)<br>3- R-DHAOx<br>4- 1rst Yescarta | R-MTX-AraC | Flubarabine + Cyclophosphamide |
| 12 | Responder | Kappa+ | 2L | 1- R-CHOP | ICE | Flubarabine + Cyclophosphamide |
| 13 | Non-responder | BCL2+, MYC+, CD30-, Higly Proliferatif (≈100%). No gene rearrangement for BCL2/BCL6/MYC. P53 overexpression | 2L | 1- R-CEOP + High Dose MTX | Ibrutinib | Bendamustine |

**Supplementary Table 2- Oncological treatment.**

Table summarising known genetic and molecular features of patient's disease, treatment line in which CAR T-cells were used, previous lines of treatment, bridging therapy and lymphodepletion regime used. Patient 9: italic treatment lines were for chronic lymphocytic leukaemia before transformation to DLBLC.

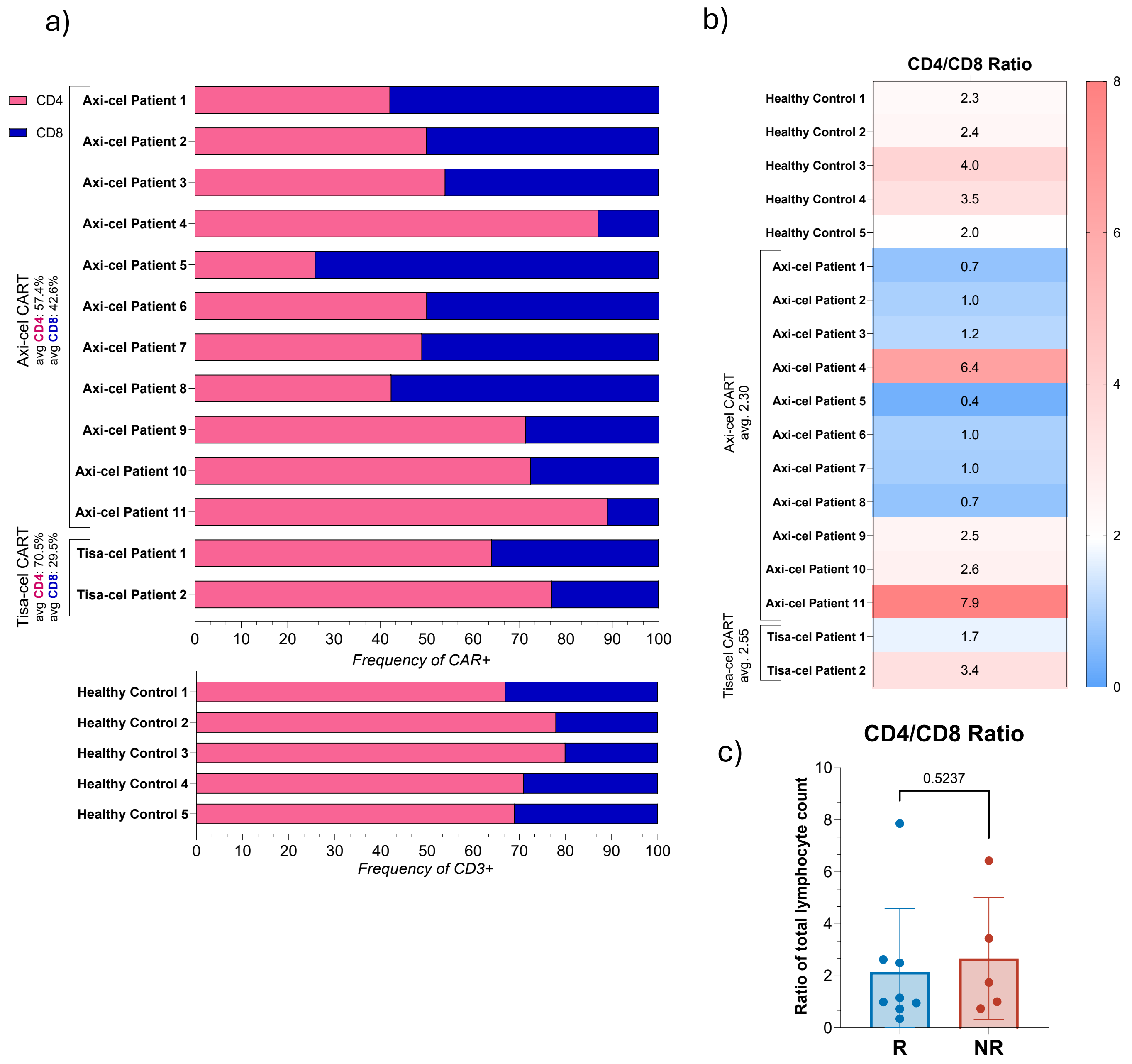

**Figure S2- Frequencies of CD4$^{+}$ and CD8$^{+}$ CAR$^{+}$ T-cells.**

Frequencies of CD4$^{+}$ and CD8$^{+}$ CAR$^{+}$ T-cells **(a)** and CD4/CD8 Ratio **(b)** in Axi-cel and Tisa-cel infusion products. This data presents the frequencies of CD4$^{+}$ and CD8$^{+}$ CAR$^{+}$ T-cells, along with the CD4/CD8 ratio, in infusion products based on CD28 (Axi-cel) and 41BB (Tisa-cel) co-stimulatory domains. These frequencies are compared to the corresponding circulating frequencies in healthy volunteers. **(c)** CD4/CD8 ratio in the infusion product of responders (R, blue) compared to non-responders (NR, red)

a)

| CAR+ T-cell Marker Frequency | CD3+ | CD3+CD4+ | CD3+CD8+ |
|---|---|---|---|
| LAG3 | 0.0031 ** | 0.0093 ** | 0.011 * |
| Ki67 | 0.05 * | 0.03 * | 0.09 ns |
| CD71 | 0.02 * | 0.02 * | 0.07 ns |
| CXCR3 | 0.35 ns | 0.60 ns | 0.02 ** |
| CXCR5 | 0.05 * | 0.17 ns | 0.05 * |
| NKG2D | 0.07 ns | 0.62 ns | 0.0031 ** |

b)

| CAR+ T-cell Marker MMI | CD3+ | CD3+CD4+ | CD3+CD8+ |
|---|---|---|---|
| CD3 | 0.03 * | 0.02 * | 0.13 ns |
| CD7 | 0.0031 ** | 0.0062 ** | 0.0016 ** |
| CD71 | 0.0192 * | 0.0295 * | 0.05 * |
| LAG3 | 0.0062 ** | 0.0062 ** | 0.02 * |
| Ki67 | 0.05 * | 0.03 * | 0.13 ns |
| CXCR3 | 0.0093 ** | 0.03 * | 0.0031 ** |
| CXCR5 | 0.05 * | 0.13 ns | 0.07 ns |
| NKG2D | 0.03 * | 0.30 ns | 0.0031 ** |

**Figure S3-Significantly expressed markers in CAR$^+$ T cells**

**(a)** Differences in frequency and **(b)** mean metal intensity (MMI) of markers between responders (blue) and non-responders (red) in CAR$^+$CD3$^+$, CAR$^+$CD3$^+$CD4$^+$, and CAR$^+$CD3$^+$CD8$^+$ compartments. Statistical significance was assessed using the Mann-Whitney U-Test, with significance levels as *$p<0.05$ and **$p<0.01$.

| Marker | Cut-off | | ROC | |
|---|---|---|---|---|
| | Sign | Value | AUC | p-value |
| CD7 | < | 114 | 0.975 | **0.0054** |
| LAG3 | > | 7.245 | 0.95 | **0.0084** |
| CXCR3 | < | 78 | 0.925 | **0.0128** |
| CD71 | > | 36.2 | 0.9 | **0.0192** |
| CD3 | < | 132.5 | 0.875 | **0.0281** |
| NKG2D | < | 13.35 | 0.875 | **0.0281** |
| CXCR5 | < | 2.41 | 0.85 | **0.0404** |
| Ki67 | > | 17.05 | 0.85 | **0.0404** |
| CD27 | < | 11.5 | 0.825 | 0.057 |
| CD4 | < | 430 | 0.7 | 0.2416 |
| CD8 | < | 52.1 | 0.7 | 0.2416 |
| CD95 | < | 54.05 | 0.7 | 0.2416 |
| TIM3 | > | 3.59 | 0.7 | 0.2416 |
| CD45RA | < | 2.13 | 0.675 | 0.3055 |
| CD45RO | < | 148 | 0.675 | 0.3055 |
| ICOS | < | 143 | 0.65 | 0.3798 |
| CD38 | < | 64.9 | 0.65 | 0.3798 |
| CD62L | < | 14.35 | 0.65 | 0.3798 |
| Perforin | < | 11.39 | 0.65 | 0.3798 |
| TIGIT | < | 11.6 | 0.625 | 0.4642 |
| CD25 | < | 307.5 | 0.6 | 0.5582 |
| CD57 | < | 50.2 | 0.6 | 0.5582 |
| CCR6 | > | 7.39 | 0.6 | 0.5582 |
| CD127 | < | 6.335 | 0.6 | 0.5582 |
| Granzyme B | > | 14.25 | 0.6 | 0.5582 |
| PD-1 | > | 9.555 | 0.575 | 0.6605 |
| BCL2 | < | 42.95 | 0.55 | 0.7697 |
| CCR4 | < | 0.0935 | 0.55 | 0.7697 |
| OX40 | > | 14.3 | 0.55 | 0.7697 |
| CD69 | > | 21.95 | 0.525 | 0.8836 |
| HLADR | > | 233.5 | 0.5 | >0.999 |
| CCR7 | > | 59.55 | 0.5 | >0.999 |

**Figure S4- MMI cutoff for response prediction.**

Summary table with power of prediction for cutfoff indicated using MMI of all markers in $CAR^{+}CD3^{+}$ T-cells with ROC curve AUC p-values and corresponding Youden index based optimal cutoff.

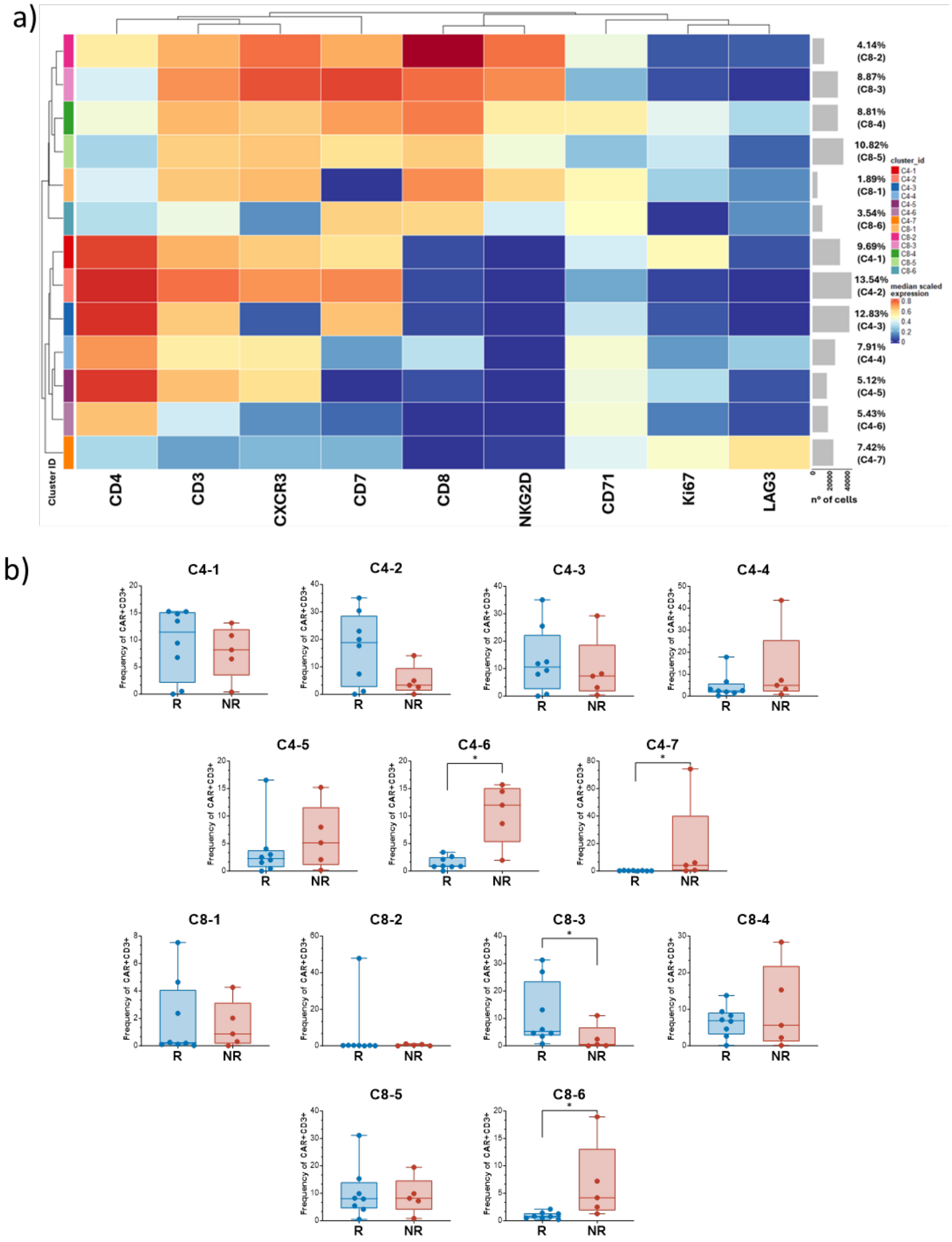

**Figure S5- UMAP analysis.**

**(a)** Unsupervised UMAP analysis using median scaled expression of markers CD3, CD4, CD8, CXCR3, CD7, NKG2D, CD71, Ki67 and LAG3 on CAR$^+$ T-cells identifies 7 CD4$^+$ (C4 1-7) and 6 CD8$^+$ (C8 1-6) distinct clusters. **(b)** Frequencies of the various clusters in the infusion product of responders (blue) and non-responders (red), expressed as frequency of CAR+ total T-cells. Statistical significance was determined using the Mann-Whitney U-Test, with significance levels indicated as *$p<0.05$.

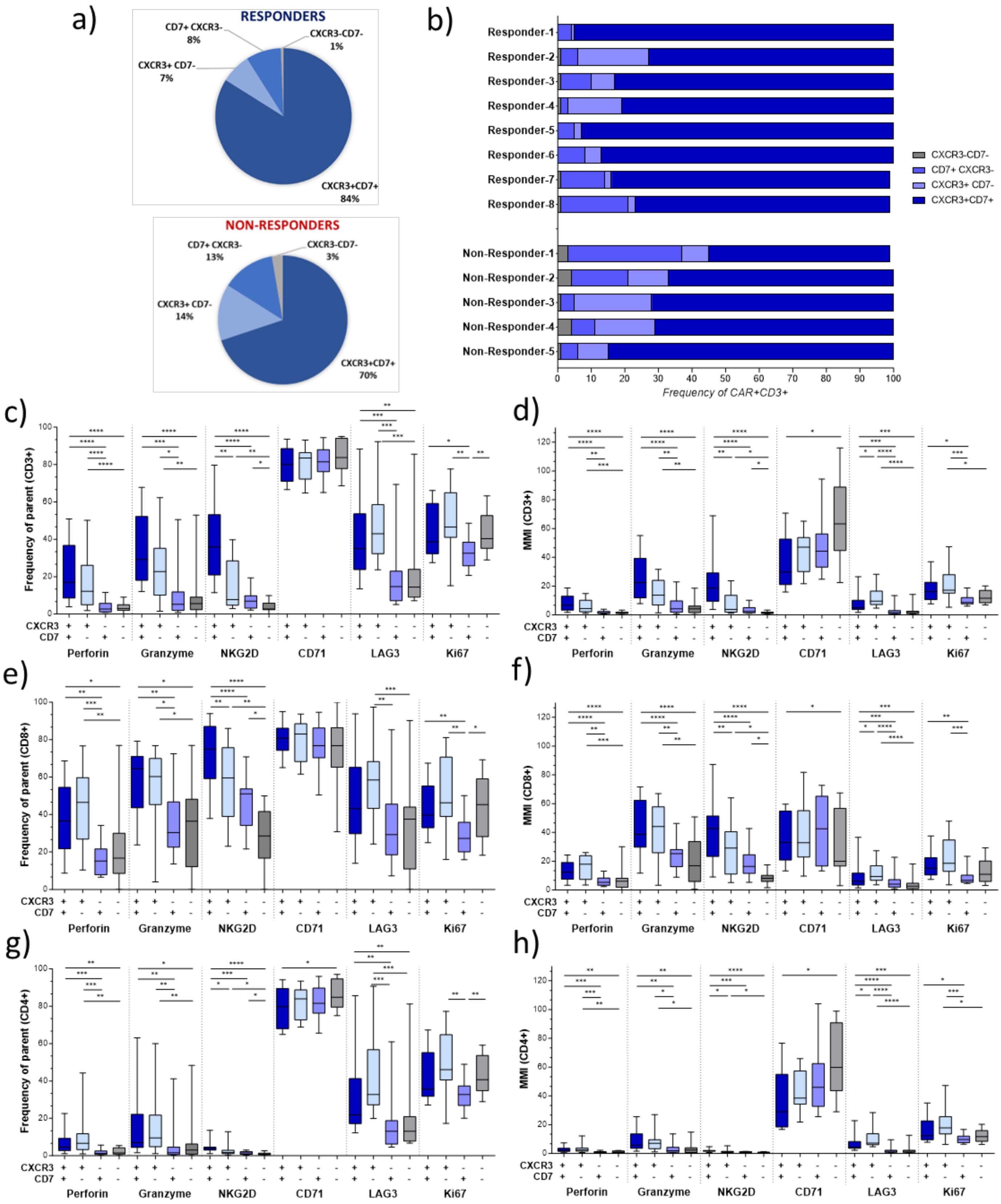
a)
RESPONDERS
CD7+ CXCR3- 8%
CXCR3-CD7- 1%
CXCR3+ CD7- 7%
CXCR3+CD7+ 84%
NON-RESPONDERS
CD7+ CXCR3- 13%
CXCR3-CD7- 3%
CXCR3+ CD7- 14%
CXCR3+CD7+ 70%
b)
Responder-1
Responder-2
Responder-3
Responder-4
Responder-5
Responder-6
Responder-7
Responder-8
Non-Responder-1
Non-Responder-2
Non-Responder-3
Non-Responder-4
Non-Responder-5
CXCR3-CD7-
CD7+ CXCR3-
CXCR3+ CD7-
CXCR3+CD7+
Frequency of CAR+CD3+
c)
Frequency of parent (CD3+)
d)
MMI (CD3+)
e)
Frequency of parent (CD8+)
f)
MMI (CD8+)
g)
Frequency of parent (CD4+)
h)
MMI (CD4+)
CXCR3
CD7
Perforin
Granzyme
NKG2D
CD71
LAG3
Ki67

**Figure S6- Distribution and marker expression of CXCR3/CD7 populations.**

**(a)**- Pie charts and **(b)** contingency table showing frequencies of 4 populations based on expression of CXCR3 and CD7. CXCR3$^+$CD7$^+$ = dark blue, CXCR3$^+$CD7$^-$ = light blue, CD7$^+$CXCR3$^-$ = blue, CXCR3$^-$CD7$^-$ = grey) in responders (n=8) and non-responders (n=5). **(b-d)** Expression of markers of interest (frequency= (**c,e,g**), MMI= (**d,f,h**) - perforin, granzyme, NKG2D, CD71, LAG3 and Ki67) in the 4 CXCR3/CD7 populations in the total CD3$^+$ population **(c-d)**, in the cytotoxic **(e-f)** (CD3$^+$CD8$^+$) and in the helper **(g-h)** (CD3$^+$CD4$^+$) compartment. Statistical significance was determined using the Mann-Whitney U Test, with significance levels indicated as *p<0.05, **p<0.01, ***p<0.001. n=13

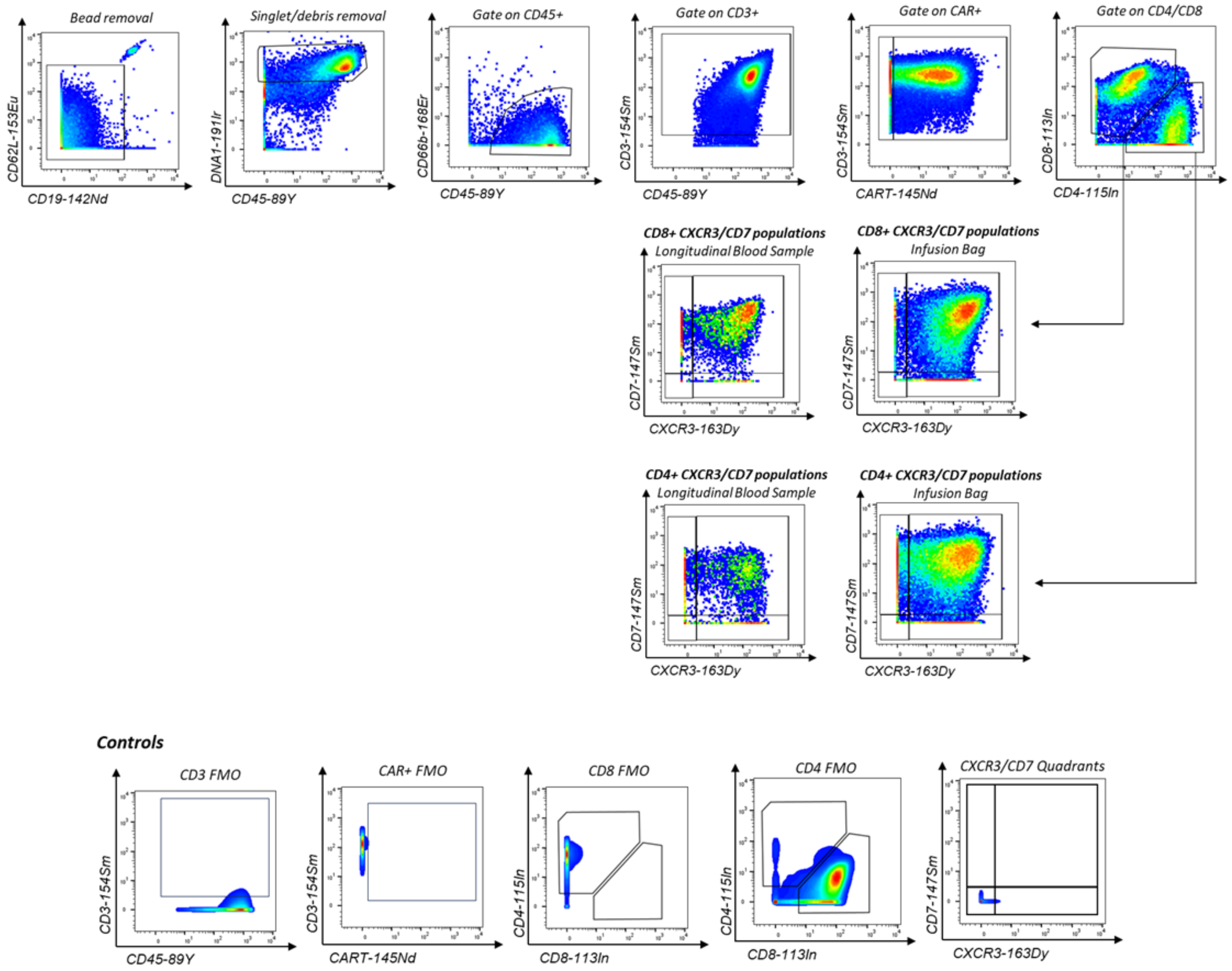

**Figure S7- Gating Strategy.**

Gating strategy used to identify CD3$^+$CAR$^+$ (both CD4 and CD8) CXCR3$^+$CD7$^+$ double positive cells. This population was identified in both infusion product and in circulation post-infusion. FMO control gates are also shown.

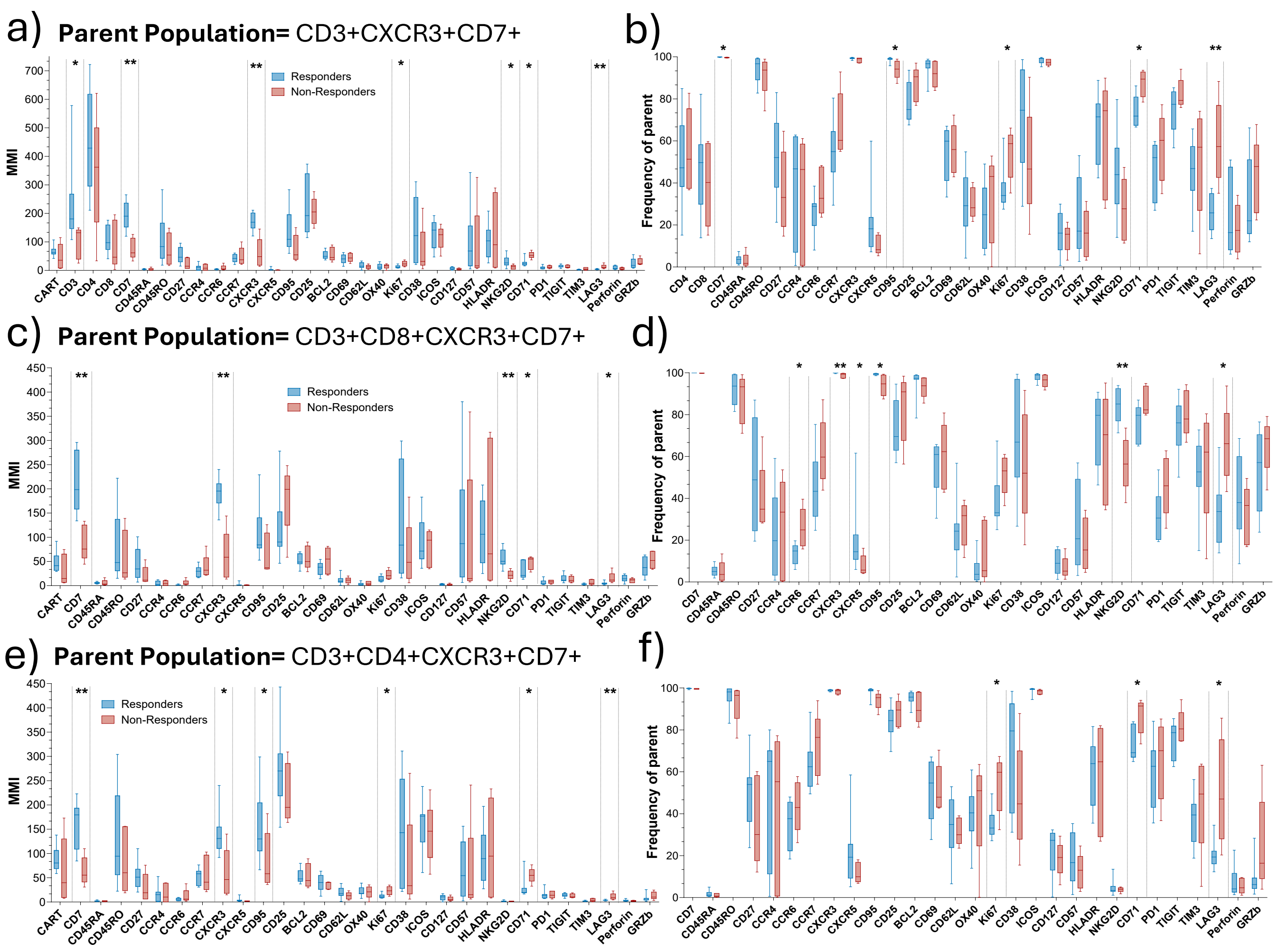

g)

| CD3 MW | p-value |
|---|---|
| CD7 | 0.00311 |
| CXCR3 | 0.00311 |
| LAG3 | 0.00311 |
| CD71 | 0.01864 |
| CD3 | 0.02176 |
| NKG2D | 0.02952 |
| Ki67 | 0.03263 |

| CD4 MW | p-value |
|---|---|
| LAG3 | 0.00311 |
| CD7 | 0.00311 |
| CXCR3 | 0.00311 |
| Ki67 | 0.01864 |
| CD71 | 0.02176 |
| CD3 | 0.02952 |
| CXCR5 | 0.11655 |

| CD8 MW | p-value |
|---|---|
| CD71 | 0.00311 |
| CXCR3 | 0.00311 |
| NKG2D | 0.00311 |
| CD71 | 0.01864 |
| LAG3 | 0.02176 |
| CXCR5 | 0.06526 |
| Ki67 | 0.06526 |

**Figure S8- Marker expression of the double positive CXCR3/CD7 populations in responders and non-responders.**

Expression of several markers as MMI **(a,c,e)** and frequency **(b,d,f)**, ranging from lineage markers, chemokine receptors, activation markers to immune-checkpoint receptors, in the double positive $CXCR3^{+}CD7^{+}$ population, comparing responders (blue, n=8) and non-responders (red, n=5). Expression of markers of the double positive population was assessed in the total $CD3^{+}$ compartment **(a-b)**, in the cytotoxic **(c-d)** ($CD3^{+}CD8^{+}$) and in the helper **(e-f)** ($CD3^{+}CD4^{+}$) compartment. **(g)** Table summarising the p-values for statistically significant markers expressed by responders and non-responders (blue= significant in responders, red= significant in non-responders). Statistical significance was determined using the Mann-Whitney U Test, with significance levels indicated as *p<0.05, **p<0.01, ***p<0.001. n=13

a)

| Population of CarT+ | Marker | Cutoff to be R | | ROC AUC | P AUC | Accuracy | PPV | NPV | Sensitivity | Specificity |
|---|---|---|---|---|---|---|---|---|---|---|
| | | Sign | Cutoff | | | | | | | |
| CD3+ | CXCR3 | > | 78 | 0.925 | 0.0128 | 0.92 | 0.89 | 1.00 | 1.00 | 0.80 |
| | CD7 | > | 114 | 0.975 | 0.0054 | 0.92 | 1.00 | 0.83 | 0.88 | 1.00 |
| | CD71 | < | 36.2 | 0.9 | 0.0192 | 0.92 | 1.00 | 0.83 | 0.88 | 1.00 |
| | NKG2D | > | 13.35 | 0.875 | 0.0281 | 0.85 | 0.88 | 0.80 | 0.88 | 0.80 |
| | LAG3 | < | 7.245 | 0.95 | 0.0084 | 0.92 | 0.89 | 1.00 | 1.00 | 0.80 |
| | Ki67 | < | 17.05 | 0.85 | 0.0404 | 0.85 | 0.88 | 0.80 | 0.88 | 0.80 |
| CD3+ CXCR3+ CD7+ | CXCR3 | > | 146 | 0.975 | 0.0054 | 0.92 | 1.00 | 0.83 | 0.88 | 1.00 |
| | CD7 | > | 133.5 | 0.975 | 0.0054 | 0.92 | 1.00 | 0.83 | 0.88 | 1.00 |
| | CD71 | < | 33.4 | 0.9 | 0.0192 | 0.92 | 1.00 | 0.83 | 0.88 | 1.00 |
| | NKG2D | > | 13.35 | 0.875 | 0.0281 | 0.77 | 0.78 | 0.75 | 0.88 | 0.60 |
| | LAG3 | < | 7.245 | 0.95 | 0.0084 | 0.92 | 0.89 | 1.00 | 1.00 | 0.80 |
| | Ki67 | < | 17.05 | 0.85 | 0.0404 | 0.77 | 0.86 | 0.67 | 0.75 | 0.80 |
| CD3+ CD4+ | CXCR3 | > | 70.15 | 0.875 | 0.0281 | 0.92 | 0.89 | 1.00 | 1.00 | 0.80 |
| | CD7 | > | 45.95 | 0.95 | 0.0084 | 0.92 | 0.89 | 1.00 | 1.00 | 0.80 |
| | CD71 | < | 37.8 | 0.875 | 0.0281 | 0.92 | 1.00 | 0.83 | 0.88 | 1.00 |
| | LAG3 | < | 4.715 | 0.95 | 0.0084 | 0.92 | 1.00 | 0.83 | 0.88 | 1.00 |
| | Ki67 | < | 16.5 | 0.875 | 0.0281 | 0.85 | 0.88 | 0.80 | 0.88 | 0.80 |
| CD3+ CD4+ CXCR3+ CD7+ | CXCR3 | > | 82.35 | 0.9 | 0.0192 | 0.92 | 0.89 | 1.00 | 1.00 | 0.80 |
| | CD7 | > | 78.55 | 0.95 | 0.0084 | 0.92 | 0.89 | 1.00 | 1.00 | 0.80 |
| | CD71 | < | 30.8 | 0.875 | 0.0281 | 0.92 | 1.00 | 0.83 | 0.88 | 1.00 |
| | LAG3 | < | 4.18 | 0.975 | 0.0054 | 0.92 | 1.00 | 0.83 | 0.88 | 1.00 |
| | Ki67 | < | 17.05 | 0.9 | 0.0192 | 0.85 | 0.88 | 0.80 | 0.88 | 0.80 |
| CD3+ CD8+ | CXCR3 | > | 151 | 0.875 | 0.0415 | 0.92 | 1.00 | 0.83 | 0.88 | 1.00 |
| | CD7 | > | 124.5 | 1 | 0.0034 | 1.00 | 1.00 | 1.00 | 1.00 | 1.00 |
| | CD71 | < | 26.35 | 0.85 | 0.0404 | 0.85 | 1.00 | 0.71 | 0.75 | 1.00 |
| | NKG2D | > | 38.65 | 0.975 | 0.0054 | 0.92 | 1.00 | 0.83 | 0.88 | 1.00 |
| | LAG3 | < | 5.94 | 0.9 | 0.0192 | 0.85 | 1.00 | 0.71 | 0.75 | 1.00 |
| | Ki67 | < | 12.85 | 0.775 | 0.107 | 0.77 | 1.00 | 0.63 | 0.63 | 1.00 |
| CD3+ CD8+ CXCR3+ CD7+ | CXCR3 | > | 154 | 0.975 | 0.0054 | 0.92 | 1.00 | 0.83 | 0.88 | 1.00 |
| | CD7 | > | 133.5 | 1 | 0.0034 | 1.00 | 1.00 | 1.00 | 1.00 | 1.00 |
| | CD71 | < | 25.25 | 0.9 | 0.0192 | 0.85 | 1.00 | 0.71 | 0.75 | 1.00 |
| | NKG2D | > | 39 | 0.975 | 0.0054 | 0.92 | 1.00 | 0.83 | 0.88 | 1.00 |
| | LAG3 | < | 5.42 | 0.9 | 0.0192 | 0.85 | 1.00 | 0.71 | 0.75 | 1.00 |
| | Ki67 | < | 12.65 | 0.825 | 0.057 | 0.77 | 1.00 | 0.63 | 0.63 | 1.00 |

b)

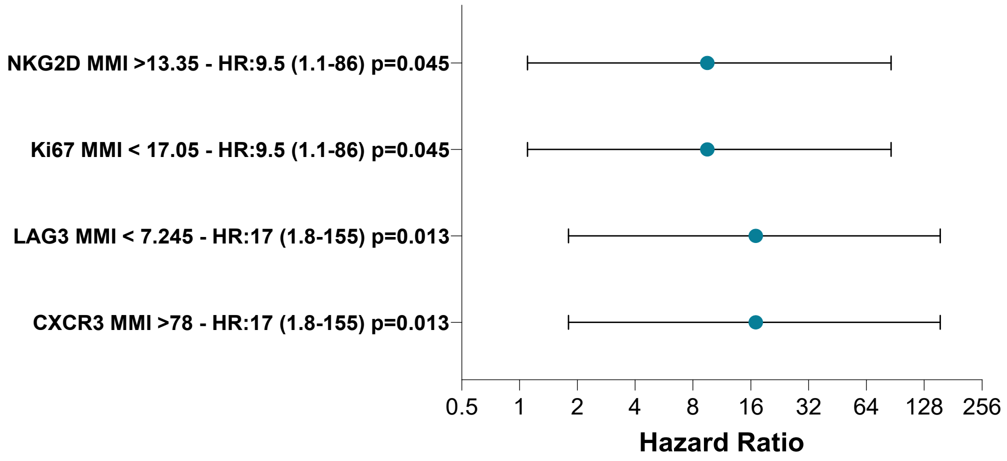

**Figure S9- Marker MMI to assess response/non-response.**

**(a)** Summary table with power of prediction for cutoff indicated using MMI of selected markers. ROC AUC and P values AUC where performed and best cutoff values optimizing the Youden index are reported. New predicted PFS accuracies, positive predictive value, negative predictive value, sensitivities and specificities have been calculated and reported in this table. **(b)** Representative COX proportional hazard ratio using Cutoff in identified in CD3+ population in the Table A.

| | | Cutoff to be R | | | | | | | | |
|---|---|---|---|---|---|---|---|---|---|---|
| Population of CarT+ | Marker | Sign | Cutoff | ROC AUC | P AUC | Accuracy | PPV | NPV | Sensitivity | Specificity |
| CD3+ | CXCR3 | > | 77.1 | 0.675 | 0.3055 | 0.77 | 0.73 | 1.00 | 1.00 | 0.40 |
| | CXCR5 | > | 16.05 | 0.85 | 0.0404 | 0.77 | 1.00 | 0.63 | 0.63 | 1.00 |
| | CD71 | < | 81.95 | 0.9 | 0.0192 | 0.85 | 0.88 | 0.80 | 0.88 | 0.80 |
| | LAG3 | < | 34.6 | 0.975 | 0.0054 | 0.92 | 1.00 | 0.83 | 0.88 | 1.00 |
| | NKG2D | > | 28.3 | 0.825 | 0.057 | 0.77 | 0.86 | 0.67 | 0.75 | 0.80 |
| | Ki67 | < | 41 | 0.85 | 0.0404 | 0.85 | 0.88 | 0.80 | 0.88 | 0.80 |
| CD3+ CXCR3+ CD7+ | CXCR5 | > | 16.8 | 0.8 | 0.079 | 0.77 | 1.00 | 0.63 | 0.63 | 1.00 |
| | CD71 | < | 82.2 | 0.9 | 0.0192 | 0.85 | 0.88 | 0.80 | 0.88 | 0.80 |
| | LAG3 | < | 43.7 | 0.95 | 0.0084 | 0.92 | 0.89 | 1.00 | 1.00 | 0.80 |
| | NKG2D | > | 49.7 | 0.775 | 0.1073 | 0.69 | 1.00 | 0.56 | 0.50 | 1.00 |
| | Ki67 | < | 45.5 | 0.85 | 0.0404 | 0.85 | 0.88 | 0.80 | 0.88 | 0.80 |
| CD3+ CD4+ | CXCR3 | > | 83.5 | 0.6 | 0.5582 | 0.69 | 0.75 | 0.60 | 0.75 | 0.60 |
| | CXCR5 | > | 15.4 | 0.75 | 0.1432 | 0.77 | 1.00 | 0.63 | 0.63 | 1.00 |
| | CD71 | < | 83.5 | 0.9 | 0.0192 | 0.85 | 0.88 | 0.80 | 0.88 | 0.80 |
| | LAG3 | < | 33.15 | 0.925 | 0.0128 | 0.92 | 0.89 | 1.00 | 1.00 | 0.80 |
| | Ki67 | < | 40.15 | 0.875 | 0.0281 | 0.85 | 0.88 | 0.80 | 0.88 | 0.80 |
| CD3+ CD4+ CXCR3+ CD7+ | CXCR5 | > | 19.75 | 0.7 | 0.2416 | 0.69 | 1.00 | 0.56 | 0.50 | 1.00 |
| | CD71 | < | 83.45 | 0.9125 | 0.0157 | 0.85 | 0.88 | 0.80 | 0.88 | 0.80 |
| | LAG3 | < | 35.05 | 0.9 | 0.0192 | 0.92 | 0.89 | 1.00 | 1.00 | 0.80 |
| | Ki67 | < | 50 | 0.875 | 0.0281 | 0.92 | 0.89 | 1.00 | 1.00 | 0.80 |
| CD3+ CD8+ | CXCR3 | > | 97.7 | 0.9 | 0.0192 | 0.85 | 1.00 | 0.71 | 0.75 | 1.00 |
| | CXCR5 | > | 8.7 | 0.85 | 0.0404 | 0.85 | 0.88 | 0.80 | 0.88 | 0.80 |
| | CD71 | < | 79.8 | 0.825 | 0.057 | 0.77 | 1.00 | 0.63 | 0.63 | 1.00 |
| | LAG3 | < | 43.55 | 0.925 | 0.0128 | 0.92 | 1.00 | 0.83 | 0.88 | 1.00 |
| | NKG2D | > | 73.15 | 0.975 | 0.0054 | 0.92 | 1.00 | 0.83 | 0.88 | 1.00 |
| | Ki67 | < | 35.05 | 0.8 | 0.079 | 0.77 | 1.00 | 0.63 | 0.63 | 1.00 |
| CD3+ CD8+ CXCR3+ CD7+ | CXCR5 | > | 9.67 | 0.875 | 0.0281 | 0.85 | 0.88 | 0.80 | 0.88 | 0.80 |
| | CD71 | < | 79.75 | 0.825 | 0.057 | 0.77 | 1.00 | 0.63 | 0.63 | 1.00 |
| | LAG3 | < | 40 | 0.925 | 0.0128 | 0.85 | 1.00 | 0.71 | 0.75 | 1.00 |
| | NKG2D | > | 74.3 | 0.975 | 0.0054 | 0.92 | 1.00 | 0.83 | 0.88 | 1.00 |
| | Ki67 | < | 48.55 | 0.825 | 0.057 | 0.85 | 0.88 | 0.80 | 0.88 | 0.80 |

**Figure S10- Marker frequencies to assess response/non-response.**

Summary table with power of prediction for cut-off indicated using frequencies of selected markers. ROC AUC and P values AUC where performed and best cutoff values optimizing the Youden index are reported. New predicted PFS accuracies, positive predictive value, negative predictive value, sensitivities, and specificities have been calculated and reported in this table.

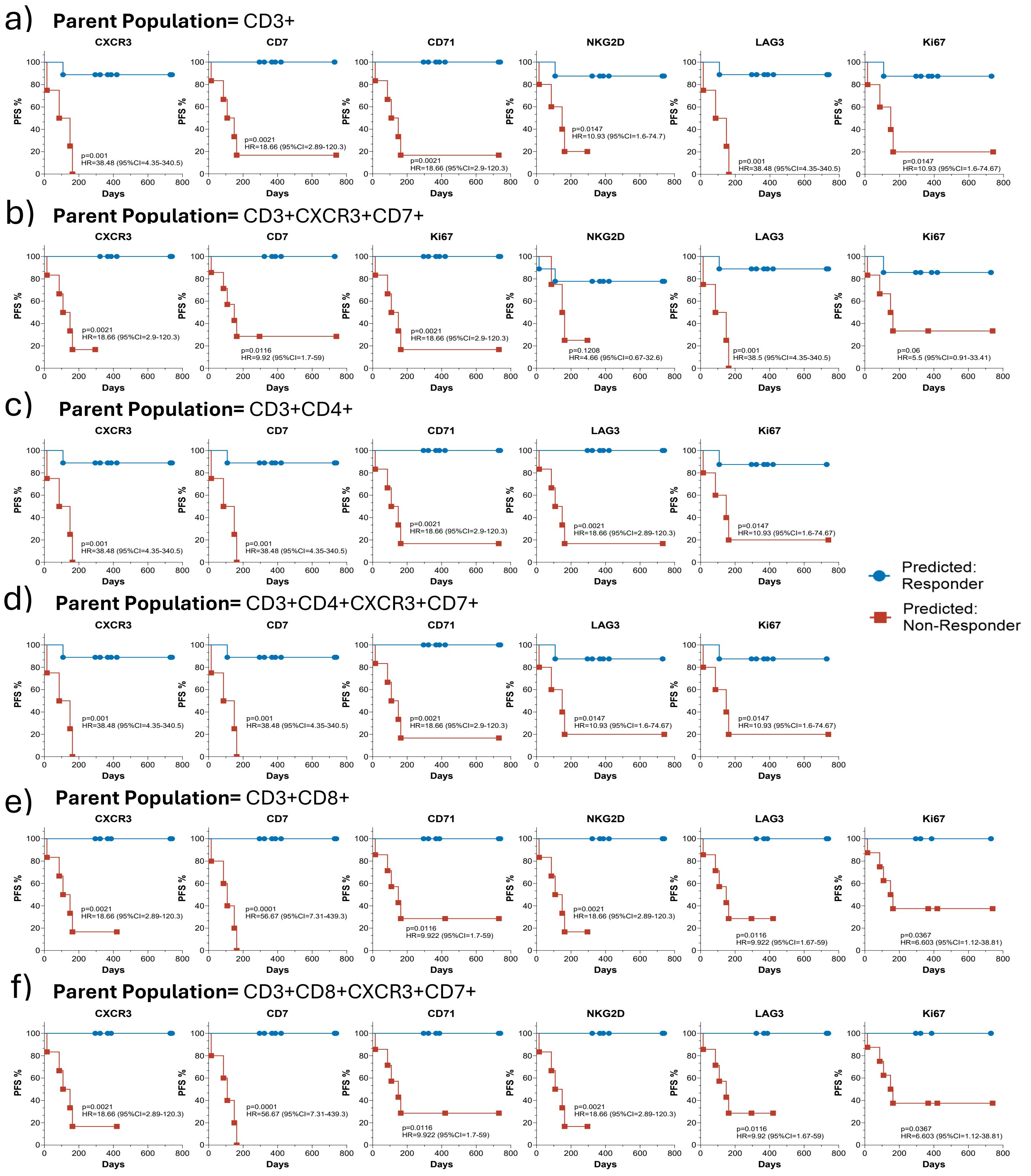

**Figure S11- Kaplan Meier curves.**

Kaplan Meier curves for 6 months PFS based on the MMI based predicted response (blue) or non-response (red) using cutoff values from Figure 3. Hazard ratios are calculated using Mante-Haenszel Method and P values using the Matel-Cox Log-rank test.

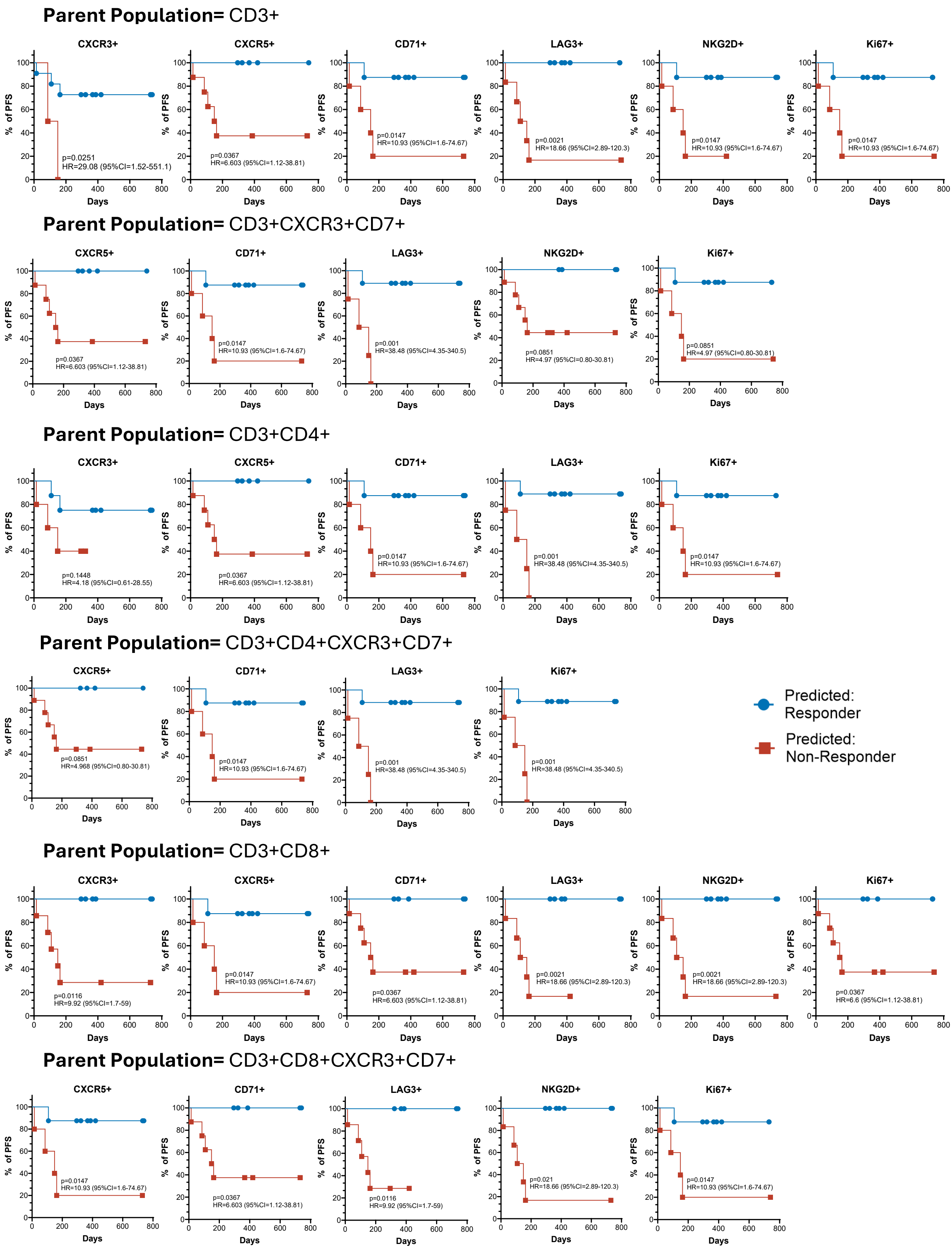

**Figure S12- Kaplan Meier curves (frequencies).**

Kaplan Meier curves for 6 months PFS based on the frequency-based predicted response (blue) or non-response (red) using cutoff values from Supplementary Figure 10. Hazard ratios are calculated using Mante-Haenszel Method and P values using the Matel-Cox Log-rank test.

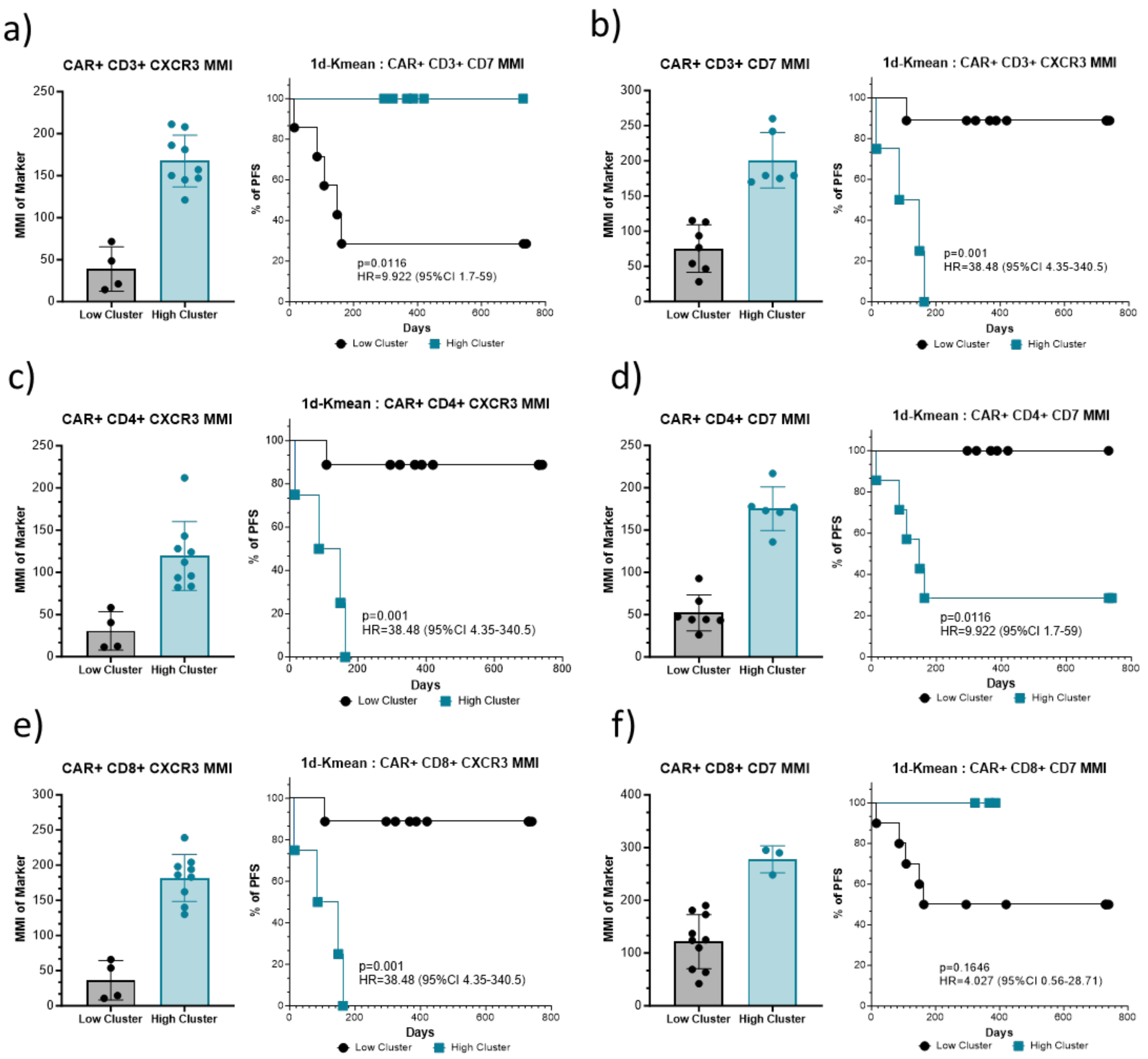

**Figure S13- PFS for low and high clusters.**

MMI of either CXCR3 (left half) or CD7 (right half) in $CD3^+$ **(a-b)**, $CD4^+$ **(c-d)**, or $CD8^+$ **(e-f)** $CAR^+$ T-cells have been used to defining 2 clusters based on the MMI levels using Kmean. Associated panels show 6 months PFS status for their respective low and high clusters (grey and blue respectively). Hazard ratios are calculated using Mante-Haenszel Method and P values using the Matel-Cox Log-rank test.

a)

| 180 Day Response | Cluster (Kmean) |
|---|---|
| **Responder** | **2** (Responders) |
| **Responder** | **2** (Responders) |
| **Responder** | **2** (Responders) |
| **Responder** | **2** (Responders) |
| **Responder** | **2** (Responders) |
| **Responder** | **2** (Responders) |
| **Responder** | **2** (Responders) |
| **Responder** | **2** (Responders) |
| **Non-Responder** | **1** (Non-Responders) |
| **Non-Responder** | **1** (Non-Responders) |
| **Non-Responder** | **1** (Non-Responders) |
| **Non-Responder** | **1** (Non-Responders) |
| **Non-Responder** | **2** (Responders) |

b)

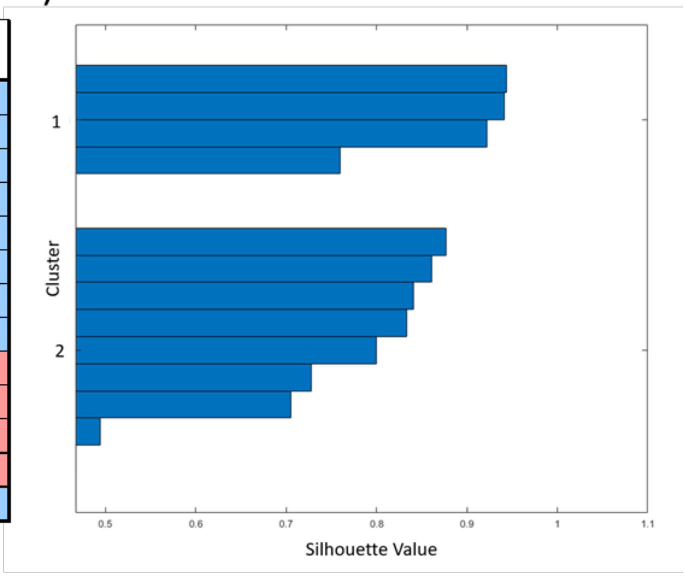

c)

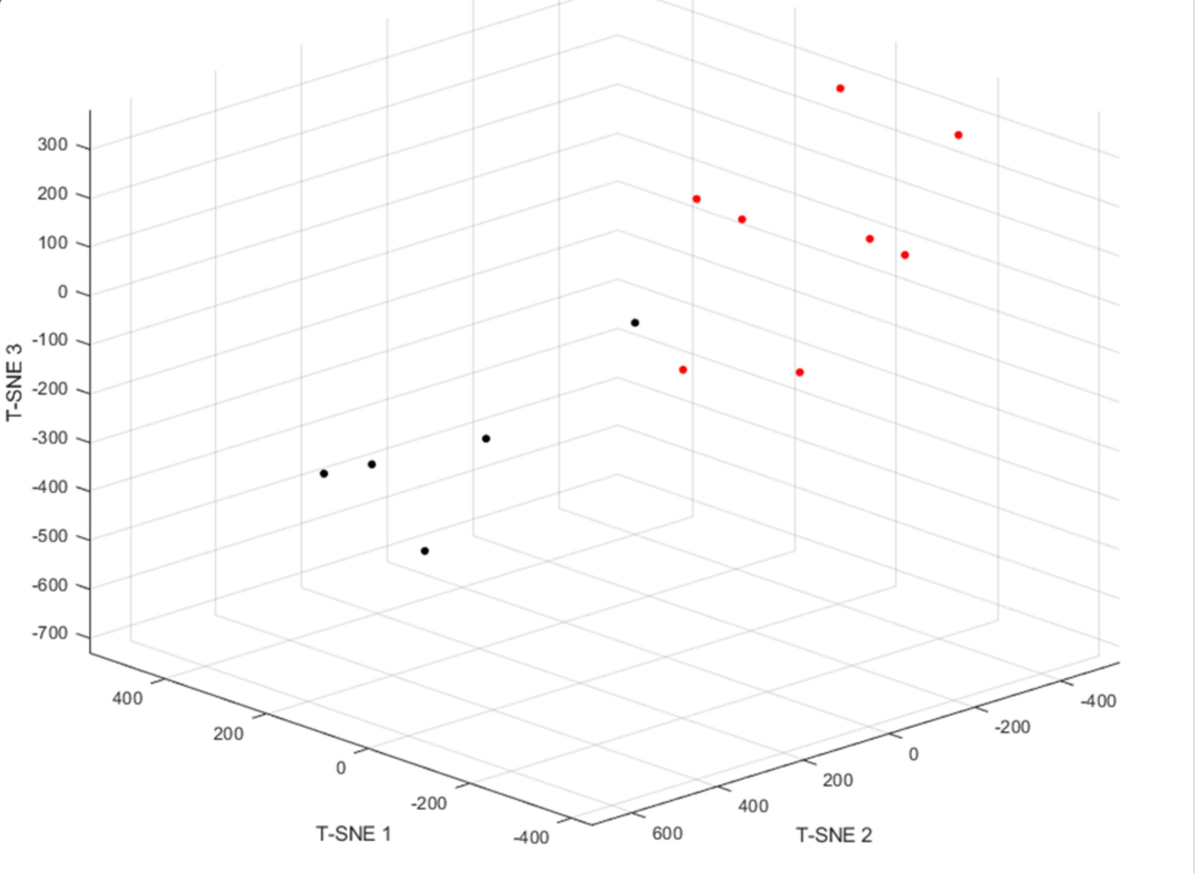

**Figure S14- K-mean clustering.**

CXCR3, CD7, CD71, NKG2D, LAG3 and Ki67 MMI in $CD3^+$ $CXCR3^+$ $CD7^+$ have been used to identify clusters using Kmean, with Predicted clusters **(a)** and associated silhouette value **(b)**. **(c)** 3D T-SNE dimension reduction representation of data with black dots showing real life 6 months non-responders and red dots responders.

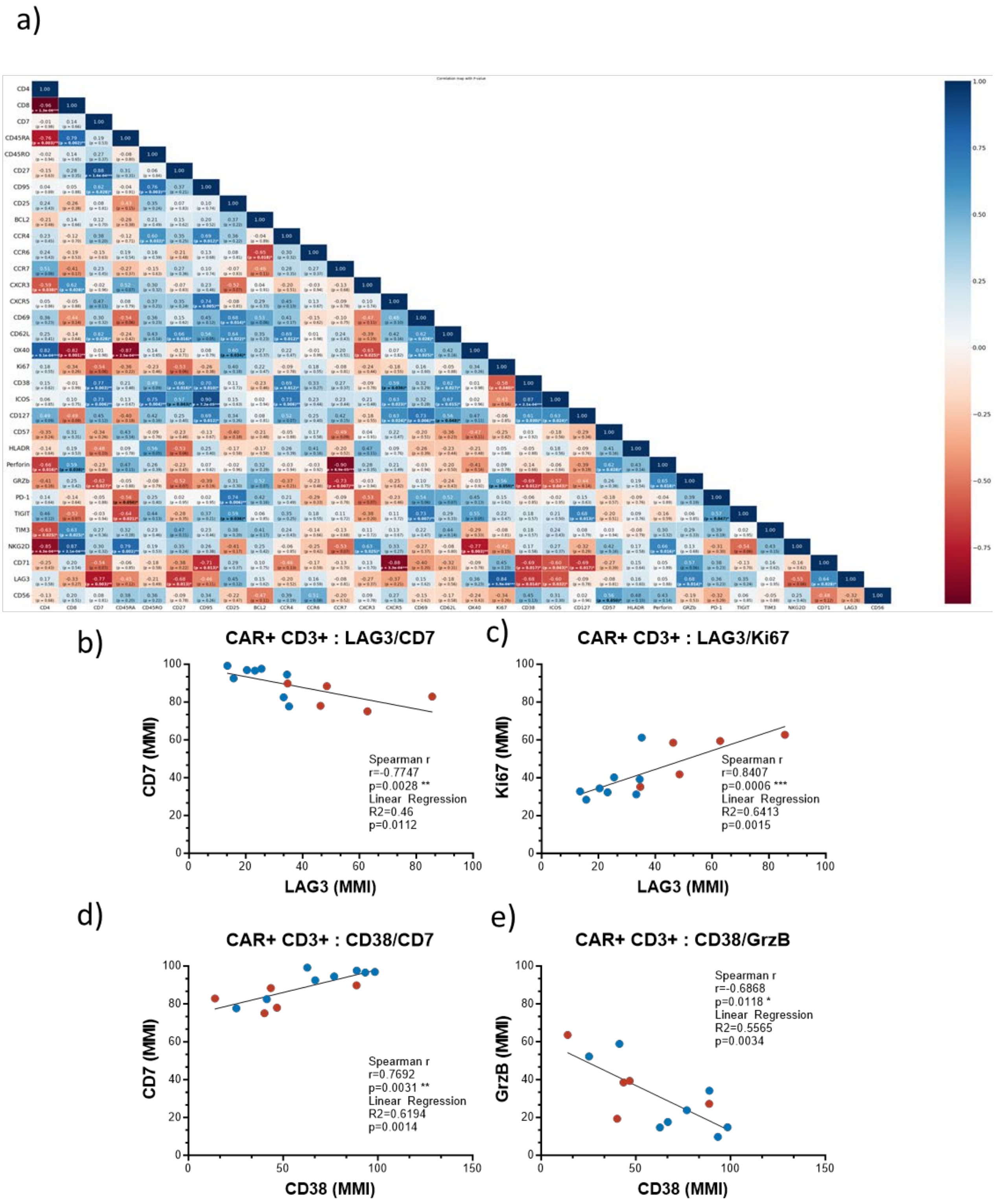

**Figure S15- All marker MMI correlation matrix (CD3).**

Correlations of MMI levels in $CAR^+$ $CD3^+$ T-cells for all markers. **(b-e)** Correlation between **(b)** LAG3/CD7, **(c)** LAG3/Ki67, **(d)** CD38/CD7, **(e)** CD38/Granzyme B MMI in $CAR^+CD4^+$ T-cells. Responders= blue, Non-Responders= red. Spearman correlation have been performed to calculate R and P values. *p<0.05, **p<0.01, ***p<0.001. Linear regression was performed and respective $R^2$ and p values are reported on the figures *p<0.05, **p<0.01, ***p<0.001

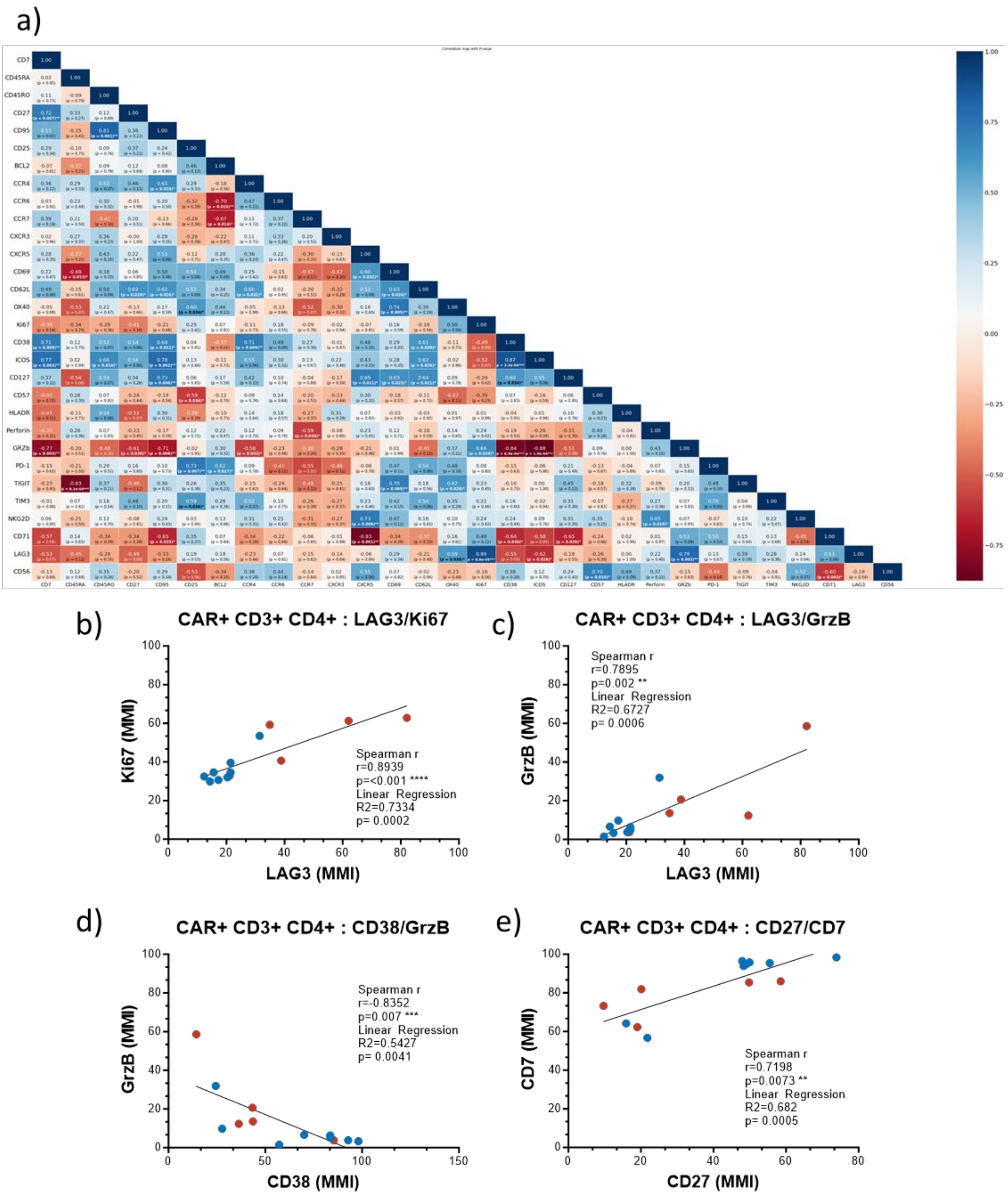

**Figure S16- All marker MMI correlation matrix (CD4).**

**(a)** Correlations of MMI levels in CAR$^+$ CD3$^+$ CD4$^+$ T-cells for all markers. **(b-e)** Correlation between **(b)** LAG3/Ki67, **(c)** LAG3/Granzyme B, **(d)** CD38/Granzyme B, **(e)** CD27/CD7 MMI in CAR$^+$CD4$^+$ T-cells. Spearman correlation have been performed to calculate R and P values. *p<0.05, **p<0.01, ***p<0.001. Linear regression was performed and respective $R^2$ and p values are reported on the figures *p<0.05, **p<0.01, ***p<0.001

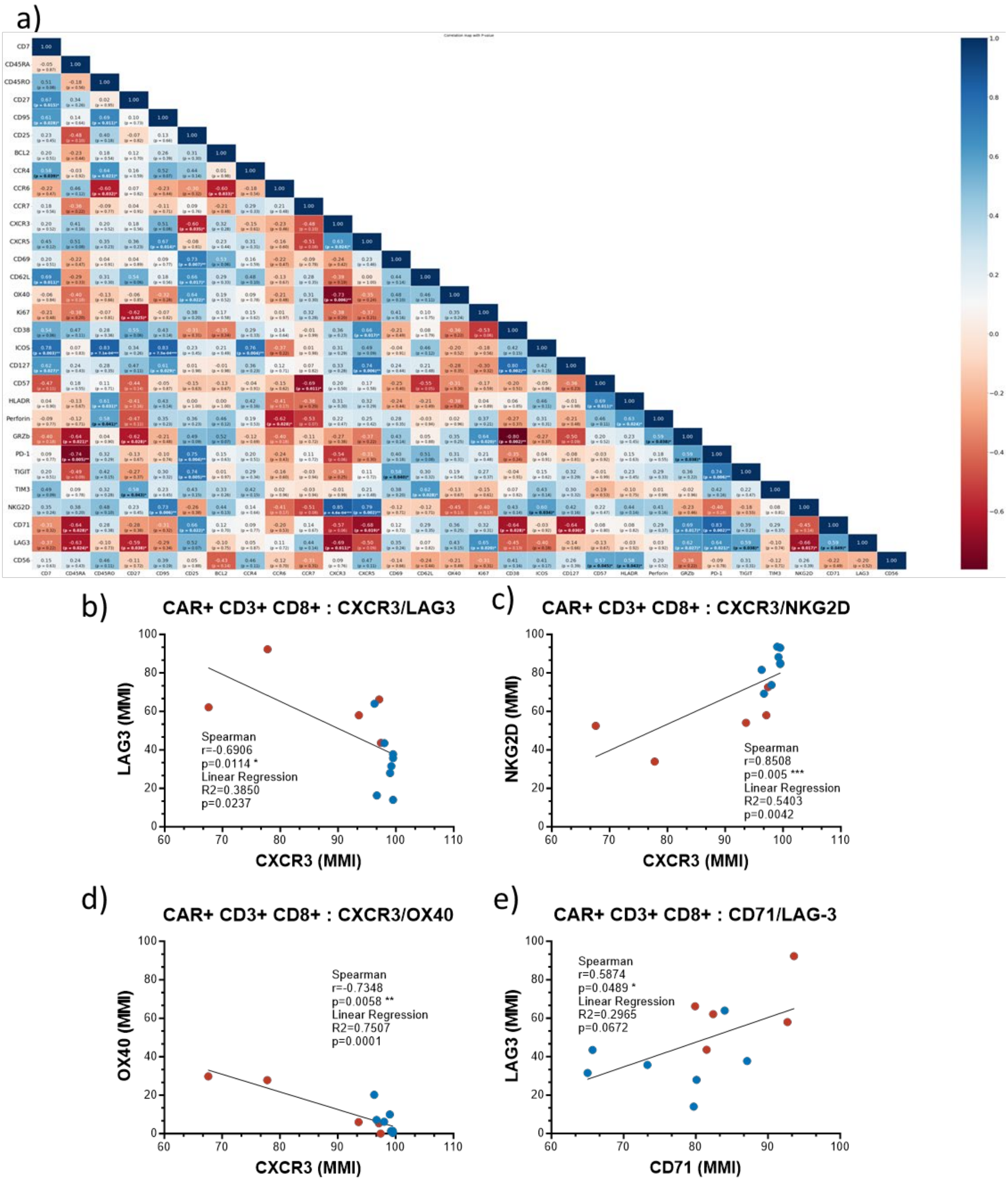

**Figure S17- All marker MMI correlation matrix (CD8).**

(a) Correlations of MMI levels in $CAR^+$ $CD3^+$ $CD8^+$ T-cells for all markers. **(b-e)** Correlation between **(b)** CXCR3/LAG3, **(c)** CXCR3/NKG2D, **(d)** CXCR3/OX40, **(e)** CD71/LAG3 MMI in $CAR^+CD8^+$ T-cells. Spearman correlation have been performed to calculate R and P values. *p<0.05, **p<0.01, ***p<0.001.

Linear regression was performed and respective $R^2$ and p values are reported on the figures $*p<0.05$, $**p<0.01$, $***p<0.001$

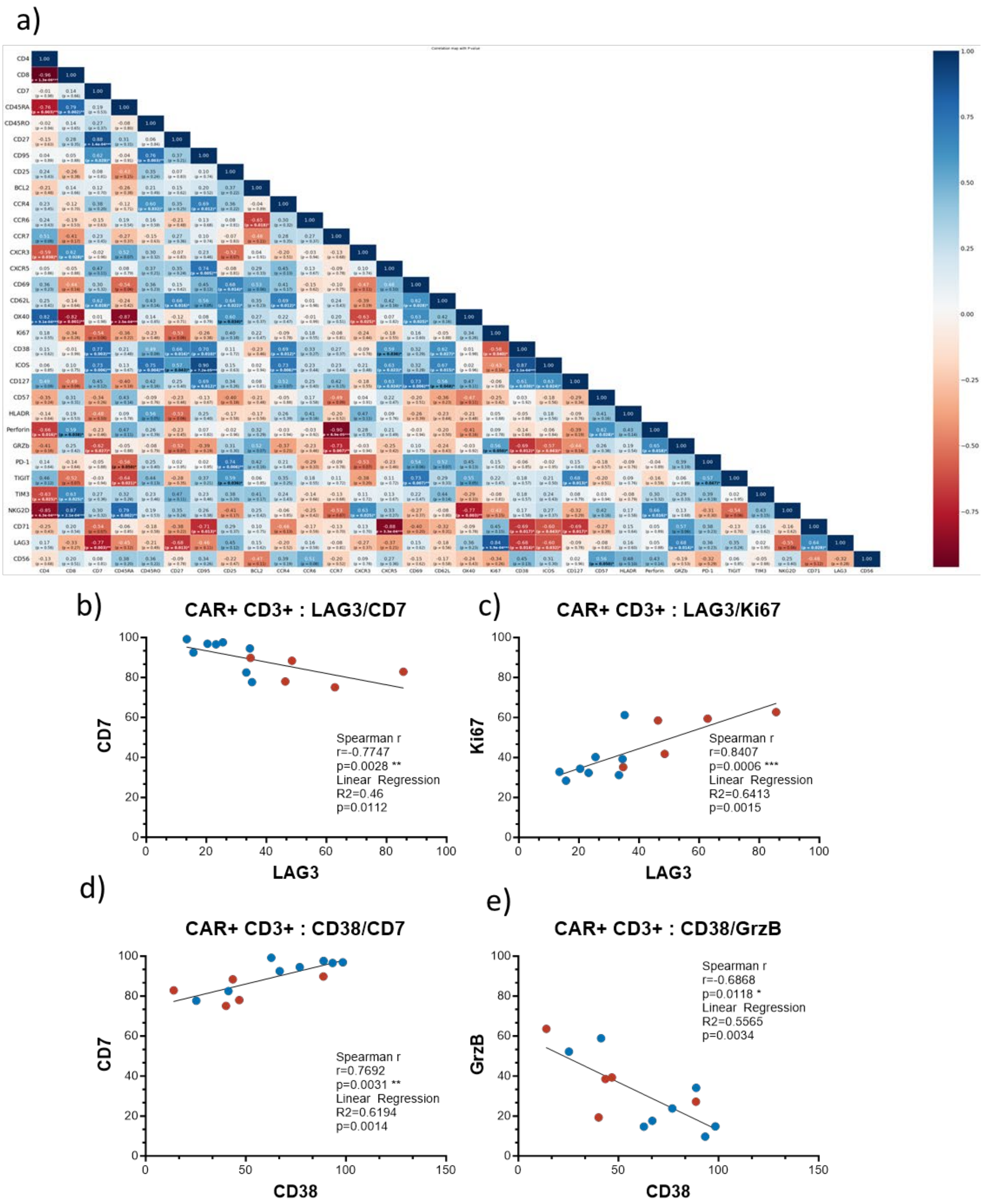

**Figure S18- All marker frequencies correlation matrix (CD3).**

**(a)** Correlations of markers frequencies levels in $CAR^+CD3^+$ T-cells for all markers. **(b-e)** Correlation between **(b)** LAG3/CD7, **(c)** LAG3/Ki67, **(d)** CD38/CD7, **(e)** CD38/GrzB frequencies in $CAR^+CD3^+$ T-cells. Spearman correlation have been performed to calculate R and P values. $*p<0.05$, $**p<0.01$, $***p<0.001$. Linear regression was performed and respective $R^2$ and p values are reported on the figures $*p<0.05$, $**p<0.01$, $***p<0.001$

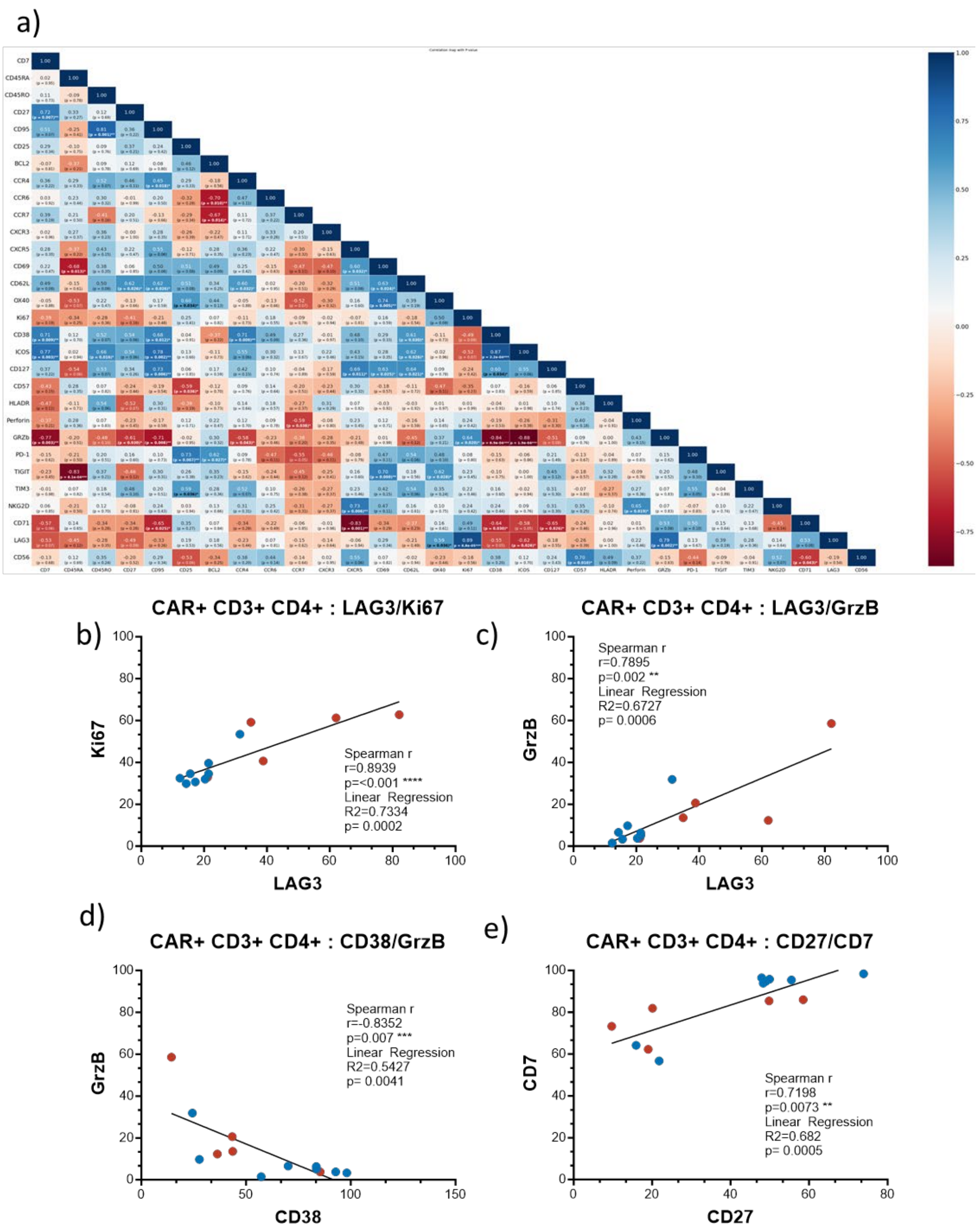

**Figure S19- All marker frequencies correlation matrix (CD4).**

Correlations of markers frequencies levels in $CAR^{+}$ $CD3^{+}$ $CD4^{+}$ T-cells for all markers. **(b-e)** Correlation between **(b)** LAG3/Ki67, **(c)** LAG3/GrzB, **(d)** CD38/GrzB, **(e)** CD27/CD7 frequencies in $CAR^{+}CD3^{+}CD4^{+}$ T-cells. Spearman correlation have been performed to calculate R and P values. *p<0.05, **p<0.01, ***p<0.001. Linear regression was performed and respective $R^2$ and p values are reported on the figures *p<0.05, **p<0.01, ***p<0.001

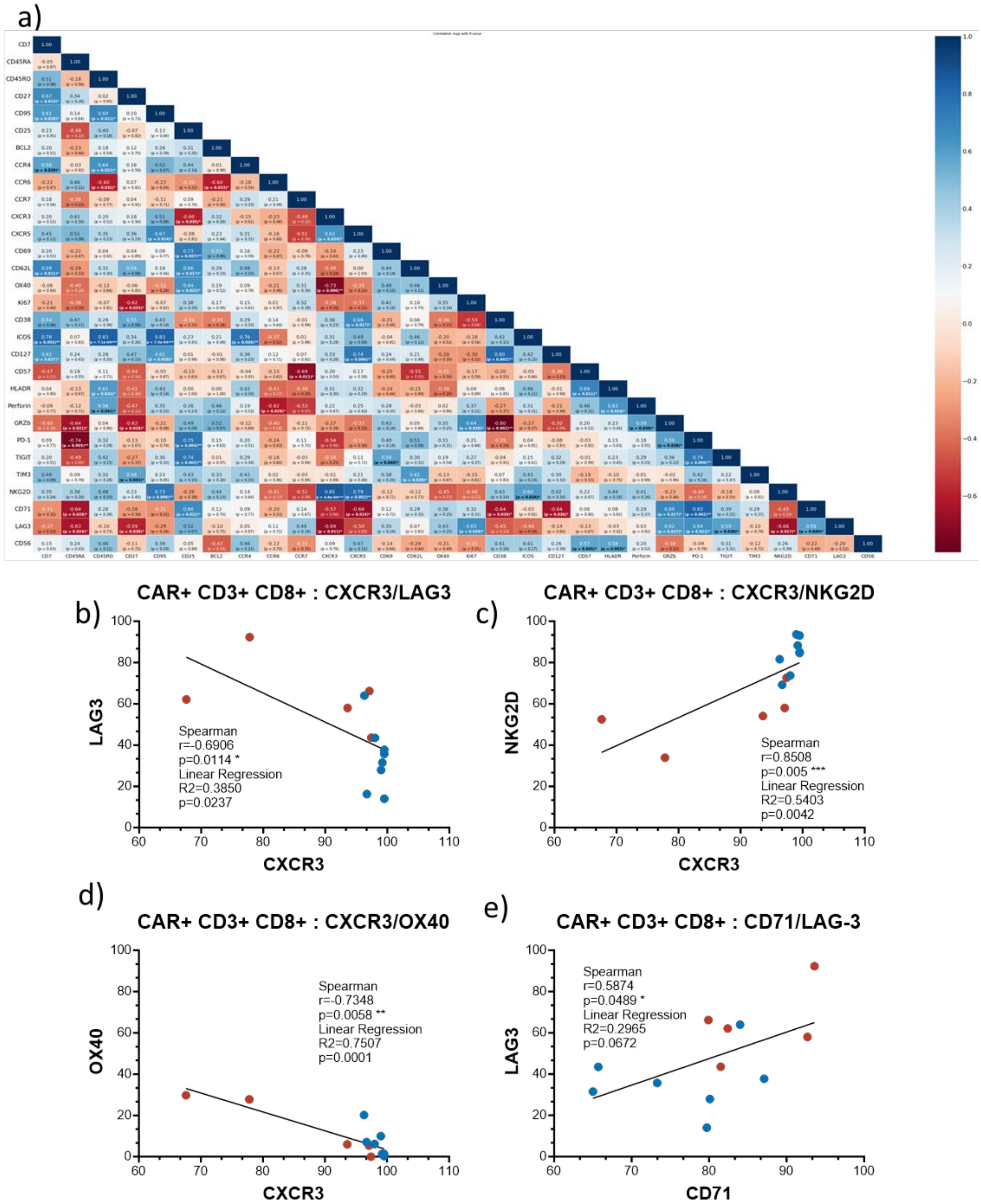

**Figure S20- All marker frequencies correlation matrix (CD8).**

**(a)** Correlations of markers frequencies levels in $CAR^+$ $CD3^+$ T-cells for all markers. **(b-e)** Correlation between **(b)** CXCR3/LAG3, **(c)** CXCR3/NKG2D, **(d)** CXCR3/OX40, **(e)** CD71/LAG3 frequencies in $CAR^+CD3^+$ T-cells. Spearman correlation have been performed to calculate R and P values. *p<0.05, **p<0.01, ***p<0.001. Linear regression was performed and respective $R^2$ and p values are reported on the figures *p<0.05, **p<0.01, ***p<0.001

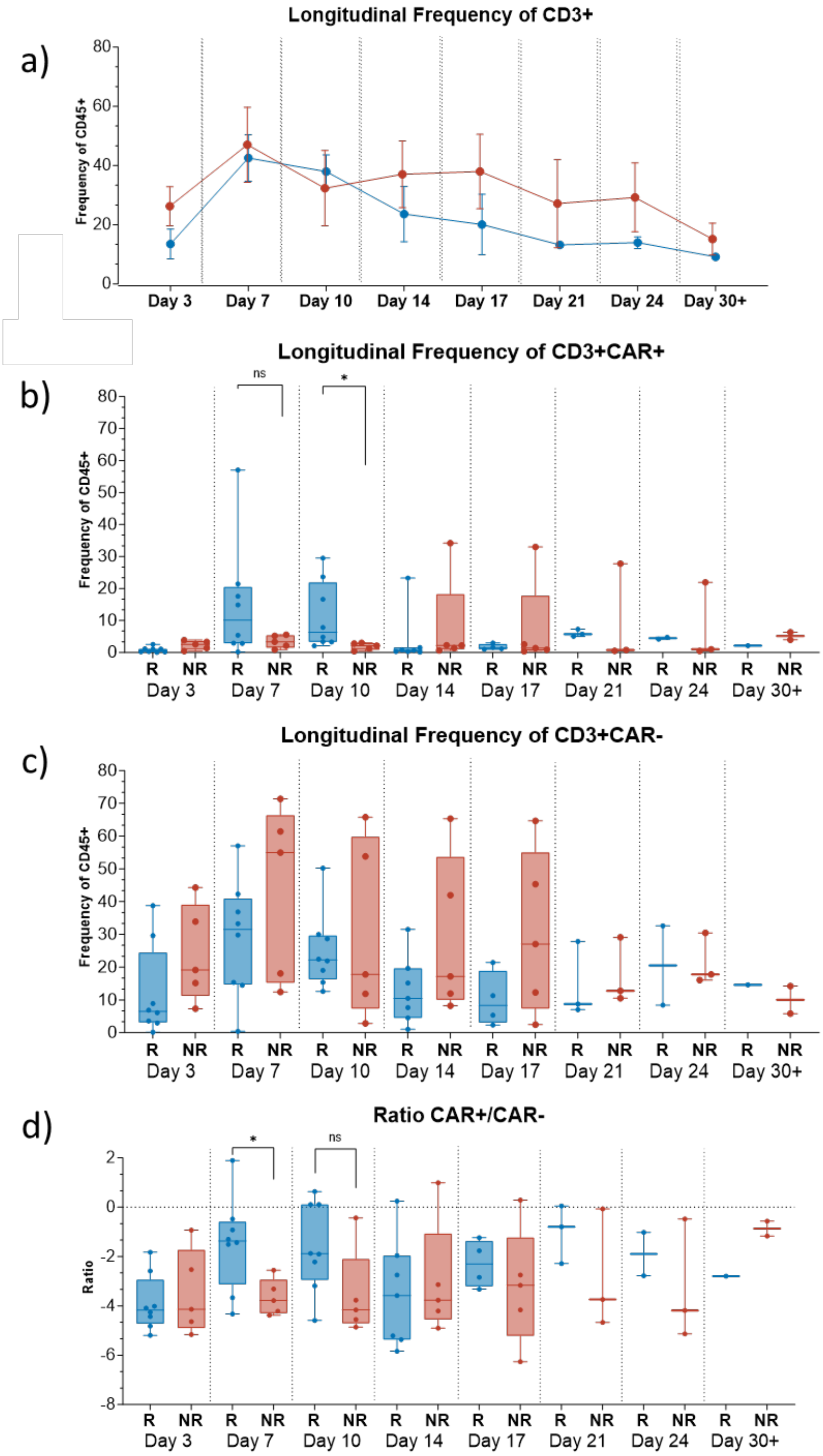

**Figure S21- Longitudinal frequencies CAR+/CAR- T-cells.**

Frequencies of $CD3^+$ cells **(a)**, $CD3^+$ $CAR^+$ T-cells **(b)**, $CD3^+$ CAR-T-cells **(c)** in the circulation of responders (blue) and non-responders (red). **(d)** Log2 of the ratio of $CAR^+$ /$CAR^-$ T-cells in R (blue) and NR (red) from Day 3 to 30 post-infusion. Statistical significance was determined using the Mann-Whitney U Test, with significance levels indicated as *p<0.05, **p<0.01, ***p<0.001.

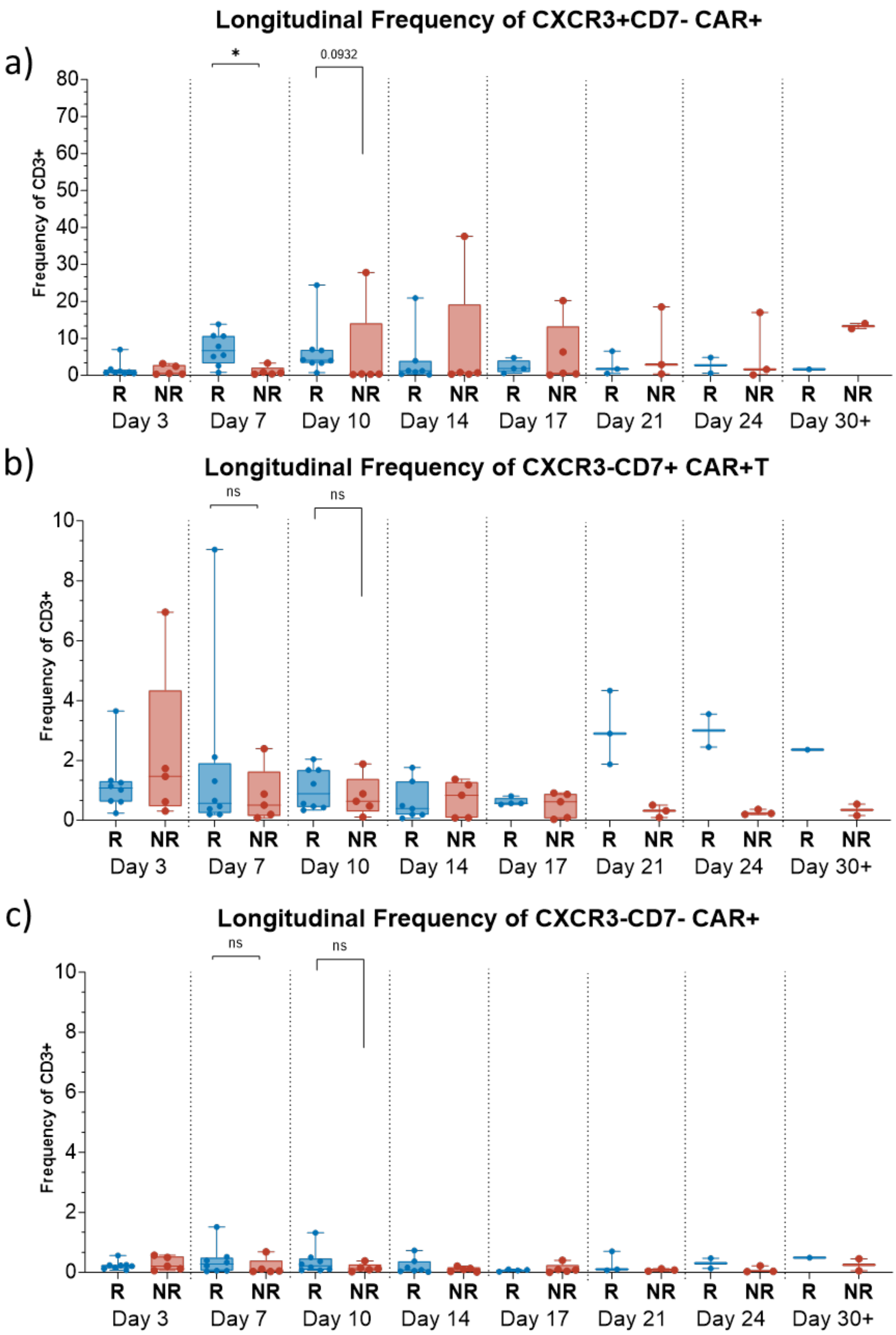

**Figure S22- Longitudinal frequencies CXCR3/CD7 populations.**

Frequencies of CXCR3$^{+}$CD7$^{-}$ **(a)**, CXCR3$^{-}$CD7$^{+}$ **(b)**, CXCR3$^{-}$CD7$^{-}$ **(c)** CAR$^{+}$ T-cells in the circulation of responders (blue) and non-responders (red) from Day 3 to Day 30 post-infusion. Statistical significance was determined using the Mann-Whitney U Test, with significance levels indicated as *$p<0.05$, **$p<0.01$, ***$p<0.001$. n=13

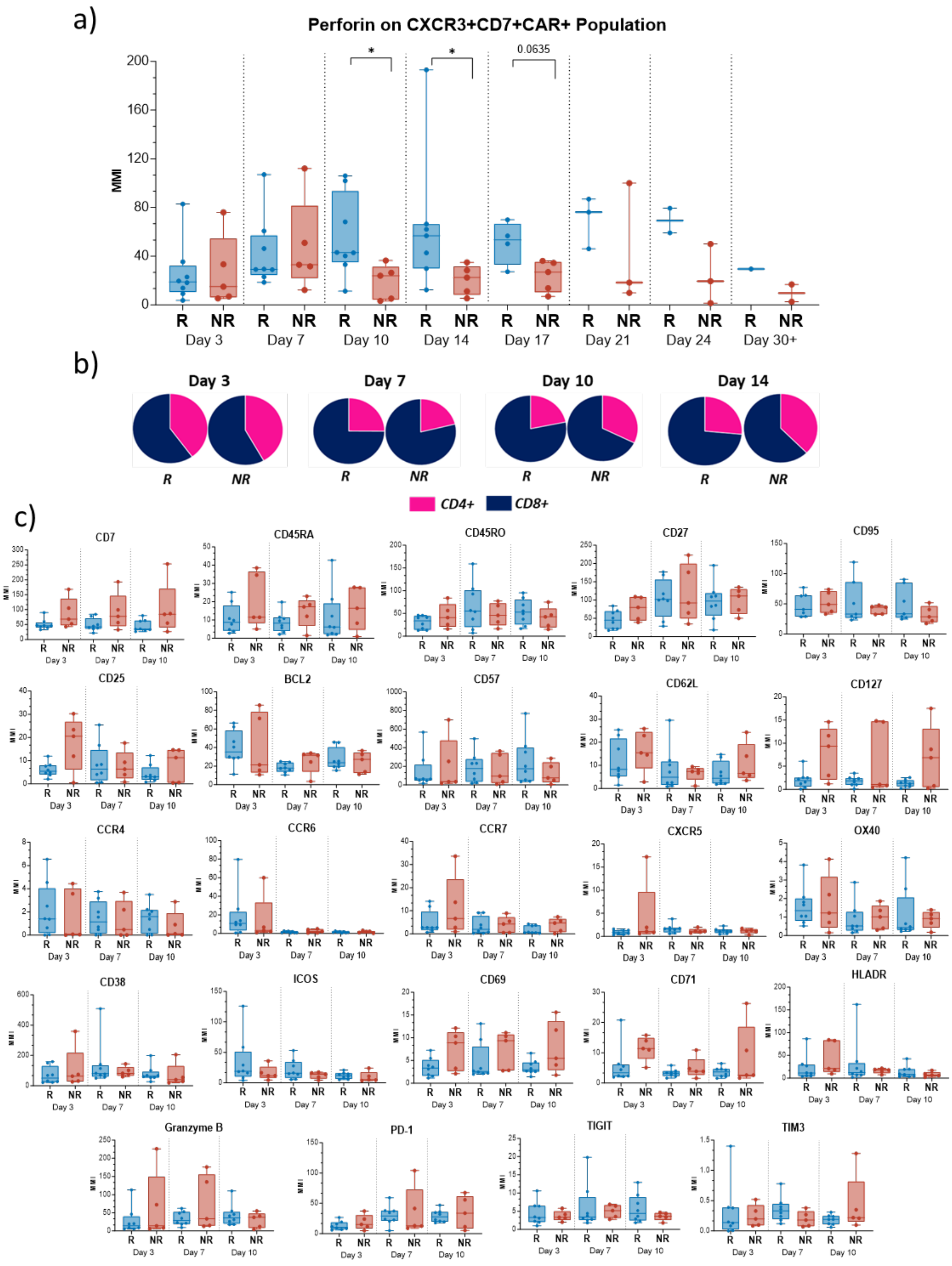

**Figure S23- Longitudinal marker expression on CXCR3$^{+}$CD7$^{+}$ CAR$^{+}$ T-cells.**

**(a)-** Expression of perforin (as MMI) on the CXCR3$^{+}$CD7$^{+}$ population from Day 3 to Day 30 post-infusion, in responders (blue) and non-responders (red). **(b)** Pie charts showing frequencies of CD4 and CD8 (of CXCR3$^{+}$CD7$^{+}$ CAR$^{+}$ T- cells) at Day 3-14 post-infusion. **(c)** Expression of all other markers (as MMI) on the $CXCR3^+CD7^+$ populations at Day 3, Day 7 and Day 10 post-infusion. Responders=blue, n=8. Non-responders= red, n=5.
